# Acute Myeloid Leukemia Skews Therapeutic WT1-specific CD8 TCR-T Cells Towards an NK-like Phenotype that Compromises Function and Persistence

**DOI:** 10.1101/2024.12.13.24318504

**Authors:** Francesco Mazziotta, Lauren E. Martin, Daniel N. Eagan, Merav Bar, Sinéad Kinsella, Kelly G. Paulson, Valentin Voillet, Miranda C. Lahman, Daniel Hunter, Thomas M. Schmitt, Natalie Duerkopp, Cecilia Yeung, Tzu-Hao Tang, Raphael Gottardo, Yuta Asano, Elise C. Wilcox, Bo Lee, Tianzi Zhang, Paolo Lopedote, Livius Penter, Catherine J Wu, Filippo Milano, Philip D. Greenberg, Aude G. Chapuis

## Abstract

Acute myeloid leukemia (AML) that is relapsed and/or refractory post-allogeneic hematopoietic cell transplantation (HCT) is usually fatal. In a prior study, we demonstrated that AML relapse in high-risk patients was prevented by post-HCT immunotherapy with Epstein-Barr virus (EBV)-specific donor CD8^+^ T cells engineered to express a high-affinity Wilms Tumor Antigen 1 (WT1)-specific T-cell receptor (TTCR- C4). However, in the present study, infusion of EBV- or Cytomegalovirus (CMV)-specific T_TCR-C4_ did not clearly improve outcomes in fifteen patients with active disease post-HCT. TCRC4-transduced EBV-specific T cells persisted longer post-transfer than CMV-specific T cells. Persisting T_TCR-C4_ skewed towards dysfunctional natural killer-like terminal differentiation, distinct from the dominant exhaustion programs reported for T-cell therapies targeting solid tumors. In one patient with active AML post-HCT, a sustained T_TCR-C4_ effector-memory profile correlated with long-term T_TCR-C4_ persistence and disease control. These findings reveal complex mechanisms underlying AML-induced T-cell dysfunction, informing future therapeutic strategies for addressing post-HCT relapse.

## Introduction

Relapsed/refractory acute myeloid leukemia (AML) after allogeneic hematopoietic cell transplantation (HCT) poses a major therapeutic challenge,^1,2^ with 2-year overall survival (OS) rate below 20% and only 4% if relapse occurs within six months post-HCT.^1–3^ Salvage therapies, including intensive chemotherapy, donor lymphocyte infusions (DLI), and second HCTs have shown limited efficacy, underscoring the urgent need for novel therapies.

The Wilms’ Tumor 1 (WT1) protein is an attractive AML immunotherapy target,^4^ as WT1 overexpression promotes proliferation and oncogenicity.^5–7^ In a phase I/II trial, targeting WT1 in the post-HCT setting, we genetically modified matched donor CD8^+^ T cells to express a high-affinity WT1-specific T cell receptor (TCRC4) specific for the HLA-A*0201-restricted WT1126-134 epitope. To minimize the potential for graft-versus-host disease (GVHD) mediated by infused donor cells,^8^ Epstein-Barr virus (EBV)-specific or cytomegalovirus (CMV)-specific (in EBV-negative donors) CD8^+^ T cells were transduced (T_TCR-C4_). Prophylactic infusion of EBV-specific T_TCR-C4_ in patients without detectable disease, but at high relapse risk post-transplant (Arm 1), yielded 100% relapse-free survival in 12 patients at a median follow-up of 44 months, and 54% in concurrent controls.^9^ However, in 15 patients with prior evidence of disease post-HCT discussed here, infused EBV-specific or CMV-specific T_TCR-C4_ did not yield a superior overall survival compared to historical controls.^1,2,10,11^

The persistence of functional antigen-specific T cells is required for sustained immunotherapy efficacy.^12–14^ In solid tumors and lymphomas, reduced persistence generally correlates with T-cell exhaustion,^12,15,16^ characterized by markers like PD-1, CTLA-4, Tim3, LAG-3, BTLA and/or TIGIT.^17^ Targeting these immune-inhibitory receptors with checkpoint-blocking antibodies mitigates exhaustion.^18,19^ In AML, recent studies have questioned the presence of T-cell exhaustion,^20^ proposing alternative mechanisms of immune dysfunction.^21,22^ Natural killer-like (NKL) markers expressed on CD8^+^ T cells have correlated with adverse outcomes, suggesting that NKL skewing may contribute to T-cell dysfunction.^22–24^ However, whether AML directly induces terminally differentiated, dysfunctional antigen-specific T cells remains unclear.^25^

Understanding the mechanisms of dysfunction that are operative in AML is critical for designing strategies to overcome the current limitations observed in T-cell therapies. Adoptive transfer of T_TCR-C4_ in refractory or relapsed AML patients has provided a unique opportunity to track AML-specific T cells, elucidate AML-induced T cell states, identify the mechanisms responsible for T-cell dysfunction, and inform the design of effective anti-AML therapies.

## Results

### T_TCR-C4_ is safe, well tolerated, and produces comparable outcomes to conventional treatments for high-risk relapsed/refractory AML patients post-HCT

From December 2012 through March 2020, 15 HLA-A2-expressing patients with relapsed and/or refractory AML post-HCT were enrolled on trial NCT01640301 (**Table 1**, **Extended data Fig. 1**). The median age at diagnosis was 40. Pre-HCT, 27% of patients had secondary or treatment-related AML, at diagnosis 47% were adverse risk, and 53% favorable/intermediate risk per European LeukemiaNet (ELN) stratification **(Supplementary Table 1)**.^26^ Post-HCT, four patients had measurable residual disease (MRD) by ∼28 days (refractory), two relapsed by 3 months, and nine relapsed after day 100. The median time to relapse was 496 days post-HCT. Thirteen patients received salvage therapy before T_TCR-C4_, including six who underwent a second HCT. Within a median of two weeks before infusion, two patients had overt disease, five were MRD-positive and the rest had no evaluable disease (NED) **(Extended data Fig. 2)**. Ten patients received EBV-specific T_TCR-C4_ and five received CMV-specific T cells (**Table 1**). Patients received one to four infusions depending on their place in the T_TCR-C4_ dose escalation **(Extended data Fig. 3)**.

**Table 1.** Clinical characteristics of AML patients receiving T_TCR-C4_ infusions. This table summarizes the clinical data of patients with AML relapsed/refractory after HCT, who received T_TCR-C4_ infusions. *AML: Acute Myeloid Leukemia, HCT: Hematopoietic Cell Transplantation, NED: No Evidence of Disease, MRD: Measurable Residual Disease, EBV: Epstein-Barr Virus, CMV: Cytomegalovirus*.

| Pt id | M/F | AML WT1 expression | Disease status 28 days post-HCT | Number of HCT before TCR-C4 infusion | Salvage therapy before TCR-C4 infusion | Lymphodepletion | Disease status at 1st TCR-C4 infusion | Days between salvage and TCR-C4 infusion | Virus specificity | TCR-C4 infusions received |
| --- | --- | --- | --- | --- | --- | --- | --- | --- | --- | --- |
| 1 | M | NA | NED | 1 | Yes | No | NED | 87 | EBV | 3 |
| 2 | F | Yes | NED | 2 | Yes | No | MRD | 60 | CMV | 4 |
| 4 | M | Yes | NED | 2 | Yes | No | NED | 89 | EBV | 2 |
| 5 | M | Yes | MRD | 1 | No | No | Overt |  | EBV | 1 |
| 6 | F | NA | NED | 2 | Yes | No | NED | 59 | CMV | 4 |
| 7 | F | No | MRD | 1 | Yes | No | MRD | 33 | EBV | 1 |
| 8 | F | Yes | MRD | 1 | No | No | MRD |  | EBV | 2 |
| 9 | F | Yes | MRD | 1 | Yes | No | MRD | 52 | CMV | 4 |
| 14 | M | Yes | NED | 2 | Yes | No | MRD | 61 | EBV | 2 |
| 15 | F | No | NED | 2 | Yes | No | NED | 110 | CMV | 2 |
| 19 | F | Yes | NED | 1 | Yes | No | Overt | 243 | EBV | 1 |
| 23 | M | Yes | NED | 1 | Yes | No | NED | 246 | EBV | 1 |
| 26 | F | Yes | NED | 1 | Yes | Yes | NED | 53 | EBV | 2 |
| 27 | F | Yes | NED | 1 | Yes | Yes | NED | 178 | EBV | 1 |
| 28 | M | Yes | NED | 2 | Yes | Yes | NED | 31 | CMV | 2 |

As in Arm 1,^9^ T_TCR-C4_ infusions were safe and well tolerated **(Supplementary Table 2)**. The incidence of acute and chronic GVHD (aGVHD, cGVHD) was lower compared to DLIs or second HCTs as salvage strategies,^11,27^ with only one patient (7%) developing grade 3 aGVHD and three (20%) developing cGVHD post-T_TCR-C4_ infusion. Post-infusion cGVHD included one grade 3, one grade 2 and one grade 1 cGVHD. However, the onset of cGVHD occurred when T_TCR-C4_ constituted <1% of the peripheral blood (PB) CD8^+^ T cells, making the infusion of T_TCR-C4_ with an endogenous virus-specific TCR the unlikely cause of cGVHD.

Among the patients enrolled (**Table 1**), four showed no overt relapse and/or MRD following TTCR- C4 infusion(s) **(Extended data Fig. 4)** supporting T_TCR-C4_’s potential biologic activity. However, when all 15 patients were analyzed together, the median OS was 242 days, the 2-year OS was 33%, and the 3-year OS was 20% **(Extended data Fig. 5)**, indicating that these individual outcomes did not translate into a significant survival advantage over historical salvage treatments.^1,2,10,11^ For example, DLI post-HCT relapse showed a 2-year OS of 21% (±3%) from relapse and 56% (±10%) from DLI in cases of remission or with a favorable karyotype, dropping to 15% (±3%) in aplasia or active disease.^27^

### Virus-specific substrate T cells and AML presence at the time of infusion are major determinants of T_TCR-C4_ persistence

In our cohort, baseline PB WT1-specific tetramer^+^ T cells were low (**Fig. 1A** – red arrows, day 0) indicating limited endogenous WT1-specific T cells. By day 28 post-infusion, four patients showed TTCR- C4 levels exceeding 3% (persistence threshold) of the CD8^+^ T cells, with three maintaining these levels beyond day 100. Higher frequencies correlated with increased absolute T_TCR-C4_ counts **(Extended data Fig. 6A**). Patients with T_TCR-C4_ frequencies below 3% were eligible for additional infusions (two infusions in 6/15 patients, >2 infusions in 4/15 patients) (**Table 1**). However, more than two infusions did not significantly increase persistence (**Fig. 1B**).

**Fig. 1.**
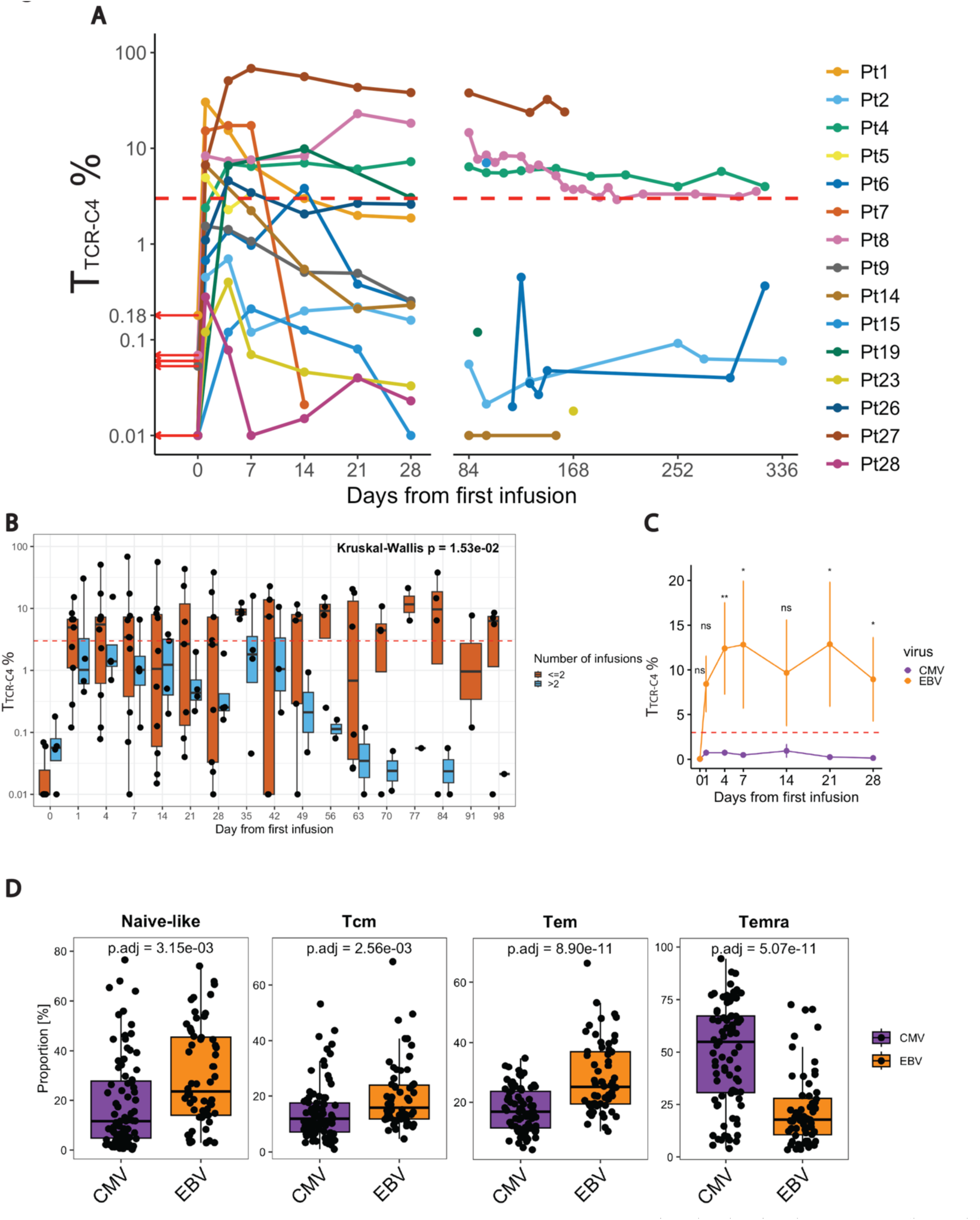
Virus and time-dependent terminal differentiation skewing of T_TCRC4_. **(A)** Line plot showing the percentage (log scale) of T_TCRC4_ in PBMCs collected after the first T_TCR-C4_ infusion for all patients (n =15), with data points representing individual samples at different timepoints post-infusion. Each patient is represented by a distinct color, and red arrows indicate the percentage of WT1-specific CD8^+^ T cells already present at day 0. **(B)** Boxplots comparing the percentage of T_TCRC4_ (log scale, y-axis) after the first infusion (x-axis), colored by the number of infusions (light blue for <= 2 infusions; dark orange > 2 infusions). The horizontal dashed red line indicates the 3% threshold used to define persisting T_TCRC4_ cells. Statistical significance was determined using Kruskal-Wallis test, with p < 0.05 considered significant. **(C)** Line plot illustrating the percentage of T_TCRC4_ (y-axis) cells derived from EBV-specific substrate cells (dark orange) or CMV-specific substrate cells (dark violet) over a 28-day period (x-axis) after the first infusion. Statistical significance was determined using Wilcoxon Rank Sum test. (* p < 0.05, ** p < 0.01) **(D)** Boxplots showing differential abundance analysis of CMV-specific (dark violet) and EBV-specific (dark orange) CD8^+^ T-cell subsets derived from the analysis of mass cytometry data.^29^ CD8^+^ T-cell subsets are defined based on marker co-expression showed in Extended data Fig. 7B. A generalized linear model was used to compute differential cluster abundance between the two conditions (CMV vs. EBV), with adjusted p-values (threshold for significance of p<0.05) displayed above each comparison.

We next explored the contribution of substrate cell virus-specificity to post-infusion persistence. Within 28 days, EBV-specific T_TCR-C4_ were more abundant than CMV-specific T_TCR-C4_, which remained below the persistence threshold (**Fig. 1C**). These differences endured when EBV-specific T_TCR-C4_ recipients from Arm 1 were included **(Extended data Fig. 6B)**.^9^ As central-memory CD8^+^ T cells (Tcm) have shown improved persistence compared to more differentiated phenotypes post-transfer,^28^ we sought to explore whether differences in endogenous virus-specific cells could recapitulate these findings. We analyzed the phenotypes of EBV- and CMV-specific T cells using mass cytometry data^29^ from 143 healthy individuals and cancer patients. Multidimensional scaling (MDS) showed segregation of CMV- and EBV-specific CD8^+^ T cells along the first dimension (MDS dim.1) suggesting distinct T-cell differentiation states between these groups **(Extended data Fig. 7A)**. Next, Flow Self-Organizing Maps (FlowSOM)^30^ metaclustering identified four CD8^+^ T-cell subsets: naïve (CD95^-^,CD45RA^+^, CCR7^+^, CD27^+^, CD28^+^), effector memory (Tem) (CD95^+^, CD45RA^-^, CD45RO^+^, CD27^+^, CD28^+^, CD127^+^), Tcm (CD95^+^, CD45RA^-^, CD45RO^+^, CCR7^+^, CD27^+^, CD28^+^, CD127^+^), CD45RA^+^ effector memory (Temra) (CD95^+^, CD45RA^+^, CD45RO^-^, CD57^+^, KLRG1^+^) **(Extended data Fig. 7B)**.^31,32^ Differential abundance analysis revealed a significant (false discovery rate/FDR < 0.05) increase of Temra cells in CMV-specific T cells and of naïve, Tcm and Tem in EBV-specific T cells (**Fig. 1D**). These results suggest that the Temra state of CMV-specific substrate cells compromises post-infusion persistence, whereas EBV-specific cells, derived from less differentiated precursors, facilitate persistence.

Persistence varied among EBV-specific T_TCR-C4_ recipients, (**Fig. 1C**, **Extended data Fig. 6A)**, with only three of ten (4, 8 and 27) exhibiting long-term persistence (>100 days post-infusion) (**Fig. 1A**, **Extended data Fig. 8A**). We reasoned that persistence was influenced by AML high-risk factors, including post-transplant remission duration and disease burden at the time of T_TCR-C4_ infusion.^3^ Patient 1 was excluded from this analysis due to unavailable WT1-expression data. Six of the remaining patients did not show long-term persistence: four (5,7,14, and 19) had detectable disease (MRD or overt) within two weeks of T_TCR-C4_ infusion, and two (23, 26) who were disease-free at infusion, had relapsed within 3 months post-HCT, suggesting difficult-to-control disease. Statistical testing revealed a trend (Fisher’s exact test, p = 0.08) linking reduced persistence with “aggressive” disease (defined as having detectable disease pre-infusion or early relapse post-HCT). Patient 8, initially categorized as “aggressive” because MRD-positive before the first infusion, became MRD-negative through salvage therapy before the second infusion, achieving the longest observed T_TCR-C4_ persistence. Reclassifying this patient as “non-aggressive”, renders the association between disease aggressiveness and persistence statistically significant (p < 0.05).These findings suggest that aggressive disease post-HCT, independently of virus-specificity, was associated with reduced T_TCR-C4_ persistence *in vivo*.

### Long-term persistent T_TCR-C4_ acquire NKL/terminal differentiation markers associated with progressive loss of function *in vivo*

To investigate the fate of T_TCR-C4_ post-transfer in patients with persistent T_TCR-C4_, and compare these with endogenous (TCR_C4_^-^) T cells, we designed a 24-color spectral flow-cytometry panel **(Supplementary Table 3)**. PB was analyzed at ∼1 (T1), ∼7 (T2), 28 (T3) days, and ∼4 months (T4) post-transfer **(Supplementary Table 4)**. FlowSOM clustering revealed five CD8^+^ T-cell states categorized into three higher-level clusters (**Fig. 2A**, **top dendrogram)**: naïve-/cm-like cells (**Fig. 2A**, **dark green)** (CD45RA^+^, CCR7^+^, CD27^+^, CD28^+^); Tem subgroup, including endogenous CD8^+^ Tem (CD28^+^, CD27^+^, Ki67^+^, CD38^+^, TIGIT^+^, PD1^+^, Tbet^+^) (**Fig. 2A**, **violet)**, and phenotypically similar T_TCR-C4__Tem (tetramer^+^) (**Fig. 2A**, **blue)**; Temra subgroup of endogenous CD8^+^ T cells (CD45RA^+^, CD57^+^, KLRG1^+^, GZMB^+^) (**Fig. 2A**, **light green)** and closely clustered T_TCR-C4__Temra (**Fig. 2A**, **red)**. Two-dimensional Uniform Manifold Approximation and Projection (UMAP) revealed a progressive increase in T_TCR-C4__Temra and decline in T_TCR-C4__Tem from T1 to T4 (**Fig. 2B**). T_TCR-C4__Temra expressed minimal Ki67 compared to T_TCR-C4__Tem (**Fig. 2A**), enabling manual gating of these two subsets (**Fig. 2C**). Over time, T_TCR-C4__Temra significantly increased, while TTCR- C4_Tem decreased (p <0.05) (**Fig. 2D**), suggesting differentiation from proliferative T_TCR-C4__Tem to non- proliferative T_TCR-C4__Temra. T_TCR-C4__Temra predominantly expressed cytotoxic/KLR markers (KLRG1, CD57, GNLY), previously linked to T-cell dysfunction in AML,^21–23^ rather than classical T-cell exhaustion markers (Tim3, PD1, and TIGIT) (**Fig. 2E**).^17,33^ Stratification by timepoint revealed progressive increase of the cytotoxic subset versus the exhausted subset (**Fig. 2F**). This phenotypic shift concided with a detectable but slow decline in T_TCR-C4_ CD8^+^ T cells producing IFNγ and TNFα (**Fig. 2G**, **Extended data Fig. 8B**). Thus, long-term persisting EBV-specific T_TCR-C4_ skew towards a distinct NKL phenotype linked to functional decline.

**Fig. 2.**
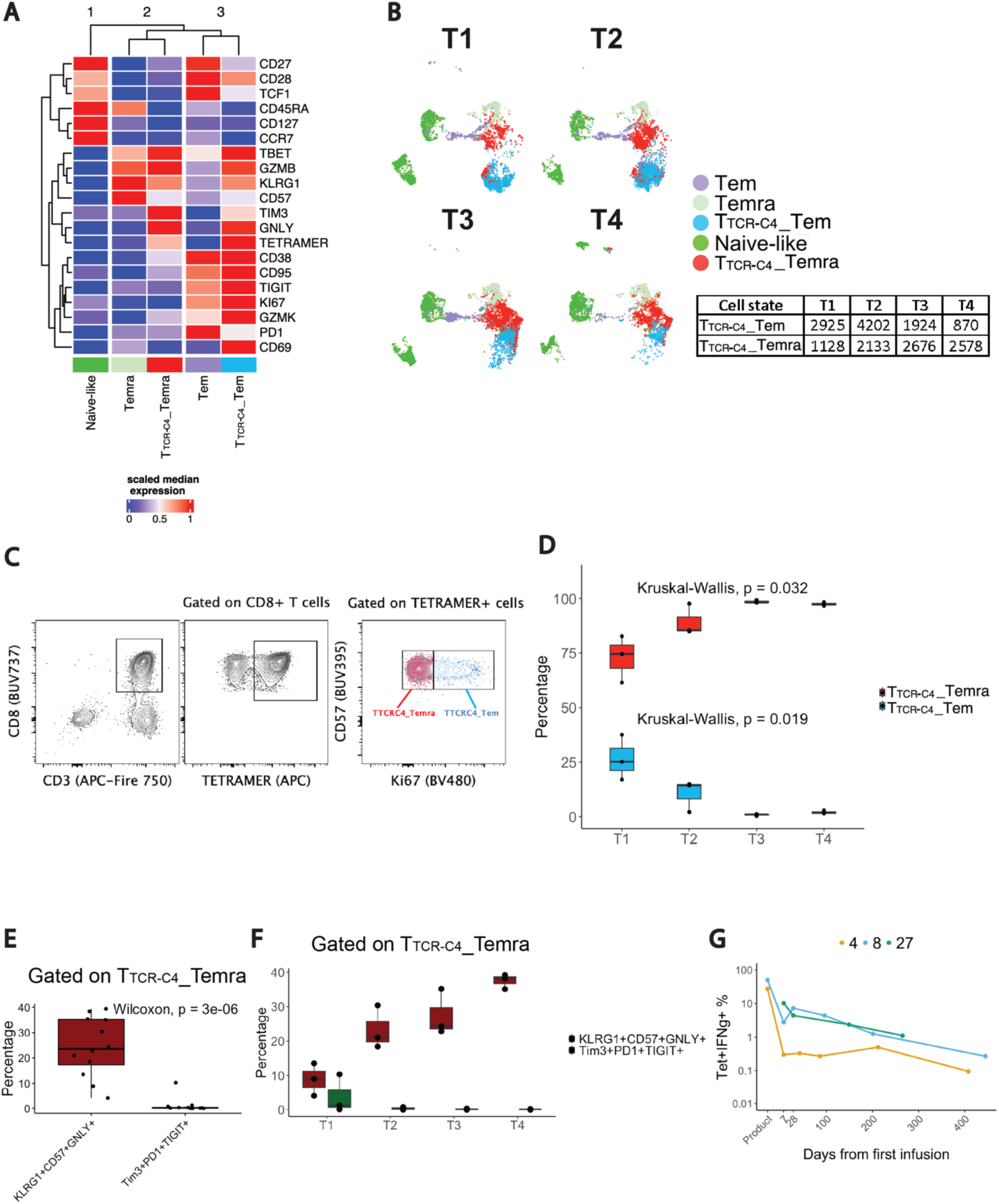
CD8^+^ T cell subset phenotypes and functional states over time. **(A)** Heatmap showing fluorescence intensity values of 20 markers across five CD8^+^ PB subsets annotated as Naïve-like (CD45RA, CCR7, CD27, CD28), Tem (CD28, CD27, Ki67, CD38, TIGIT, PD1, Tbet), T_TCRC4__Tem (TETRAMER, CD28, Ki67, CD38, TIGIT, GZMB, Tbet, KLRG1, GNLY), Temra (CD45RA, CD57, KLRG1, GZMB, Tbet), T_TCRC4__Temra (TETRAMER, GNLY, KLRG1, TIM3, GZMB, Tbet). The median marker expression identifies the markers that characterize each subset with red indicating high expression and blue low. The data were scaled after aggregation to highlight population-level differences between tetramer^+^ and tetramer^-^ samples, ensuring a clear visualization of marker expression patterns across subsets. Hierarchical clustering on the left dendrogram groups markers based on their expression patterns, revealing co-occurring markers across subsets, while the top dendrogram shows relationships between subsets based on their marker profiles, revealing similarities and differences. The K-means algorithm was applied to further group the clusters by similarity into three groups marked by the numbers (1,2,3) at the top of the heatmap. Data were derived from spectral flow- cytometry analysis. **(B)** UMAP plots showing the two-dimensional distribution of the annotated CD8^+^ T-cell subsets, with the plots colored according to the subset annotation and split by timepoint. The table shows the absolute numbers of T_TCR-C4__Tem and TTCR- C4_Temra cells at each timepoint (T1, T2, T3, T4). **(C)** Contour plots illustrating the gating strategy for T_TCR-C4__Tem (blue) and T_TCR-C4__Temra (red). This strategy starts by identifying the CD3^+^CD8^+^ cell population, followed by selecting tetramer^+^ CD8^+^ cells. Cells are then gated based on Ki67 expression, with Ki67^-^ cells (red) classified as T_TCR-C4__Temra and Ki67^+^ cells (blue) as TTCR- C4_Tem. This gating strategy is based on the marker expression shown in the heatmap in Fig. 2A. **(D)** Boxplots depicting the percentage (y-axis) of T_TCRC4__Tem and T_TCRC4__Temra populations identified in Fig. 2C over time (x-axis, from T1 to T4). Statistical significance was assessed using Kruskal-Wallis test (threshold for significance: p< 0.05). **(E)** Boxplot showing the percentage (y- axis) of cytotoxic KLRG1^+^, CD57^+^, GNLY^+^ (dark red) compared to exhausted TIM3^+^, PD1^+^, TIGIT^+^ (dark green) T cells among T_TCRC4__Temra. Statistical significance was assessed using the Wilcoxon rank-sum test, with a significance threshold of p < 0.05. **(F)** Boxplots displaying the percentage (y-axis) of cytotoxic (dark red) compared to exhausted (dark green) T cells among T_TCRC4__Temra over time (x-axis, T1 to T4). **(G)** Line plot showing the TTCR_C4_^+^IFNg^+^ cells following WT1-peptide stimulation. The x axis represents the days from the first infusion, while the y axis indicates the percentage of tetramer^+^IFNg^+^ cells (log10 scale) gated on CD8^+^ T cells.

### T_TCR-C4_ largely represent an intermediate state between effector memory and NK-like/terminally differentiated cells

To explore the relationship between CD8^+^ endogenous T-cell and T_TCR-C4_ states, we performed single-cell RNA sequencing (scRNAseq) on available PB and bone marrow (BM) samples from patients with T_TCR-C4_ exceeding 3% of CD8^+^ T cells at least until day 28 post-infusion. This analysis included two prophylactic^9^ and five treatment-arm patients **(Supplementary Table 5)**.

Unsupervised clustering of PB CD8^+^ T cells (n = 24,472) identified 13 clusters, with the TCRC4 transgene expressed across several clusters, but primarily in cluster 2 (**Fig. 3A**). Using the marker-based purification algorithm scGate,^34^ we identified TCR_C4_^+^ cells (**Fig. 3B**) labeled as T_TCR-C4_, while endogenous (TCRC4^−^) T cells included clusters 5 and 6,labeled as naïve-/cm-like, expressing *CCR7*, *SELL*, *TCF7*, *LEF1*, *IL7R*; clusters 2, 4 and 11, labeled as Tem, expressing genes associated with activation (*CD69*, *TIGIT*, *GZMK*); cluster 12, labeled as interferon signaling genes (ISG), expressing *ISG15*, *ISG20*, *IRF7*, *IFI6*; clusters 9 and 10, labeled as T memory/proliferative (Tmem/prolif), expressing proliferation (*MKI67*, *MCM5*, *MCM7*) and memory (*CD27*) genes; clusters 0,1,3,7, and 8, labeled as NKL/Temra, expressing genes associated with cytotoxicity/NKL (*KLRF1*, *KIR3DL1*, *NKG7*, *FCGR3A*, *GZMB*, *KLRD1*, *PRF1*) (Fig. 3A-C, Extended data Fig. 9A-C, Supplementary Table 6).

**Fig. 3.**
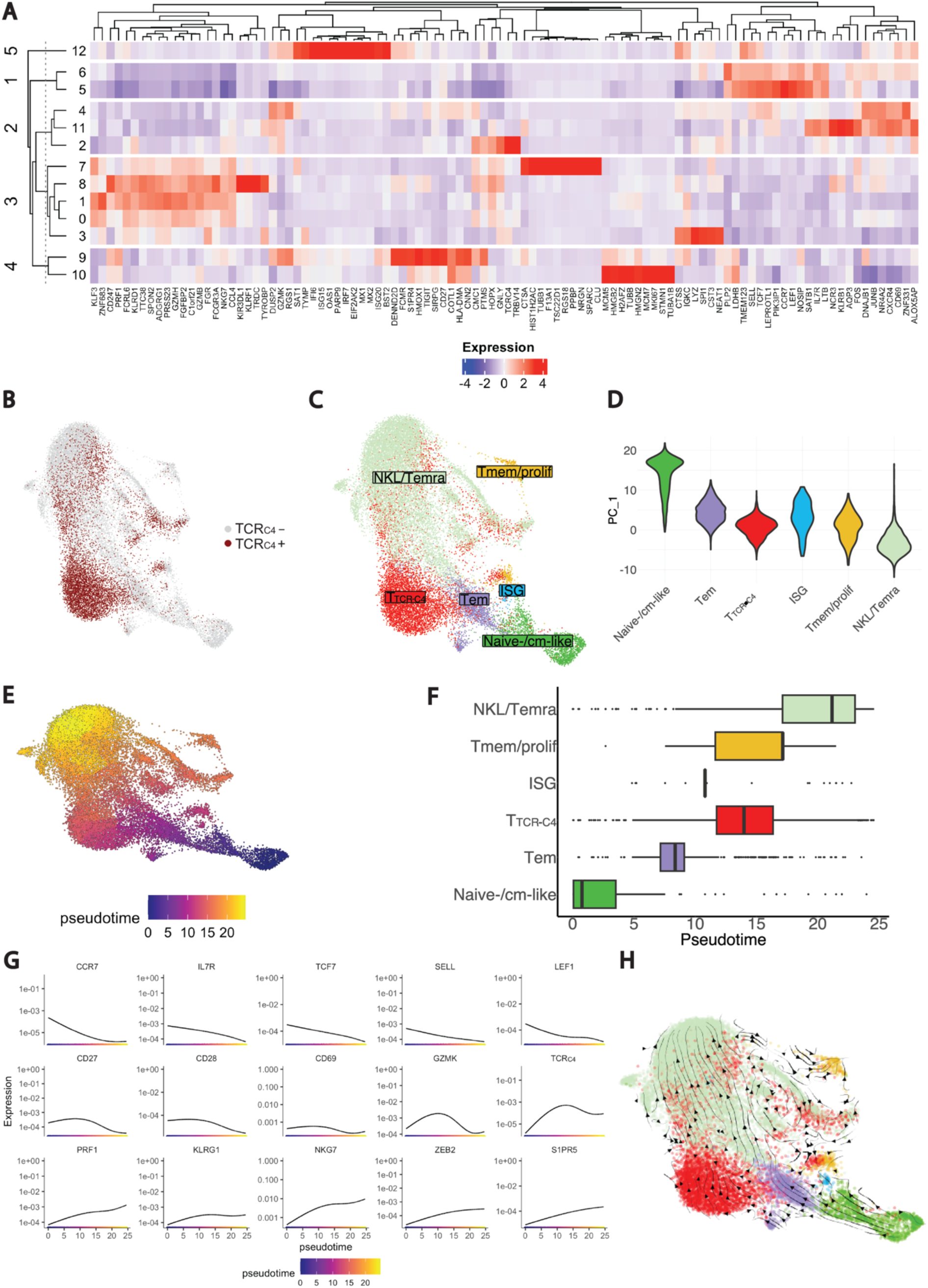
Single cell transcriptomic analysis of endogenous, T_TCRC4_ CD8^+^ T-cell states and their differentiation dynamics. **(A)** Heatmap showing the top 10 differentially expressed genes identified within each cluster. The “top 10” refers to the 10 genes with the most significant differential expression across the identified clusters. The dendrogram on the left displays the similarity between the 13 clusters, which were determined through unsupervised clustering based on gene co-expression patterns. The top dendrogram shows the relationships between the genes based on their expression patterns. To further refine this clusters, a K-means algorithm was applied to group the 13 clusters into 5 main categories, labeled 1 to 5 on the left side of the heatmap. Blue represents low gene expression, while red indicates high gene expression. **(B)** UMAP plot illustrating the distribution of CD8^+^ T cells (endogenous and T_TCR-C4_^+^) on a two-dimensional space. T_TCR-C4_^+^ cells were identified using scGate,^34^ an R package which scores cells based on TCR_C4_ expression and defines thresholds to classify cells as positive or negative for the population of interest. TCR_C4_^+^ cells are colored in dark red, while TCR_C4_^-^ cells (endogenous) are represented in light grey. **(C)** UMAP plot displaying the two-dimensional distribution of annotated CD8^+^ T-cell transcriptional states, colored by subset. **(D)** Violin plot illustrating the distribution of CD8^+^ T-cell subsets identified through gene expression (Fig. 3A) and TCR_C4_ score (Fig. 3B) along the principal component 1 (PC_1) axis. Each violin represents a different CD8^+^ T-cell state. The proximity of each subset along PC_1 (y-axis) indicates transcriptional similarity, with subsets closer together showing more similar gene expression profiles **(E)** UMAP plot illustrating the inferred developmental trajectory of CD8^+^ T-cell transcriptional states, as predicted by Monocle.^79^ The color scale, ranging from blu to orange/yellow, represents the pseudotime, where blue indicates earlier developmental stages and orange/yellow indicates later stages of the trajectory. **(F)** Boxplot showing the distribution of the 5 CD8^+^ T-cell subsets (y-axis) along the pseudotime (x-axis). The position of the boxes along the pseudotime axis provides context for how the subsets are positioned within the developmental trajectory, with subsets at lower pseudotime values representing earlier stages of differentiation, while those at higher pseudotime values correspond to later stages. **(G)** Line plots displaying the smoothed gene expression of selected genes characterizing the 5 CD8^+^ T-cell subsets along the pseudotime. These plots illustrate the dynamic changes in gene expression (y-axis) as cells progress along the inferred trajectory (x-axis). The smoothed curves show how the expression of each gene varies at different pseudotime points. **(H)** UMAP plot colored by the annotated CD8^+^ subsets overlaid with the predicted velocity stream computed through scVelo.^41^ The velocity streams represent the predicted direction and magnitude of gene expression changes for each individual cell, providing insights into the dynamic transitions between cellular states. These streams highlight the likely trajectories cells follow as they evolve over time, offering a predictive view of future cellular states based on current transcriptional dynamics.

To assess transcriptional similarities among cell states, we performed principal component analysis, positioning naïve-/cm-like and NKL/Temra as differentiation spectrum extremes, with T_TCR-C4_ as the intermediate state between these two extremes (**Fig. 3C**).

To ensure unbiased labeling of CD8^+^ T cells, we constructed a scRNAseq reference atlas comprising 109,051 CD8^+^ T cells, using a published scRNAseq dataset of tumor-infiltrating lymphocytes.^35^ We aligned and projected our scRNAseq dataset onto the reference atlas **(Extended data Fig. 9B)**, confirming the correspondence between the transcriptional states in our dataset and the reference. Differential gene expression (DGE) analysis using manually curated markers further supported this annotation **(Supplementary Table 7, Extended data Fig. 9C)**.^31,32,36,37^

Consistent with previous work,^32^ we observed a progressive reduction of stem-like markers (*IL7R*, *TCF7*) from naïve/cm-like cells through Tem and NKL/Temra cells, while activation markers (*CD69*, *GZMK*) peaked in Tem. NKL markers, including KLR-exhaustion markers (*S1PR5*, *ZEB2*),^38,39^ were highest in NKL/Temra. T_TCR-C4_ expressed activation and NKL markers, reflecting an intermediate state between Tem and NKL/Temra (**Extended data Fig. 9D**).

Monocle^40^ trajectory inference confirmed T_TCR-C4_ as an intermediate state between Tem and the terminal NKL/Temra state (**Fig. 3E-F**), and removing the TCRC4 transgene did not alter this differentiation trajectory **(Extended data Fig. 10A-B).** Single-gene expression over pseudotime revealed early peaks of naïve/stem-like markers (*CCR7*, *IL7R*, *TCF7*, *SELL*, *LEF1*) (**Fig. 3G**, **first row**), naïve/memory (*CD27*, *CD28*) and activation markers (*GZMK*, *CD69*), with the latter remaining high throughout mid-pseudotime before declining.(**Fig. 3G**, **second row)**. TCRC4 expression peaked at mid-pseudotime, but remained expressed until the end of pseudotime. NKL/Temra-associated genes (*PRF1*, *KLRG1*, *NKG7*, *ZEB2*, *S1PR5*) peaked at the end of pseudotime (**Fig. 3G**, **third row)**. RNA velocity inference^41^ supported the spectrum of differentiation from naïve-like to Tem and T_TCR-C4_, culminating in NKL/Temra cells (**Fig. 3H**).

Overall, our findings suggest that T_TCR-C4_ largely existed in an intermediate state between the Tem phenotype and the NKL/Temra stage.

### AML drives T_TCR-C4_ towards NKL/terminal differentiation instead of the dominant exhaustion pattern associated with T cells in solid tumors

To investigate AML’s impact on the differentiation of endogenous CD8^+^ T cells and T_TCR-C4_, patients samples were grouped as AML- (no AML detected, including two prophylactic arm cases),^9^ or AML+ (blasts evident in BM and/or PB) **(Supplementary Table 8)**. The increase of ISG and NKL/Temra in AML+ and T_TCR-C4_ in AML- (**Fig. 4A**) was significant (**Fig. 4B**), with a wide confidence interval for ISG, making the magnitude of this difference unclear. T_TCR-C4_ overexpressed NKL/Temra genes in AML+ compared to AML- (**Fig. 4C**), suggesting that AML blasts may induce a transcriptional shift towards NKL/Temra in T_TCR-C4_ and endogenous T cells.

**Fig. 4.**
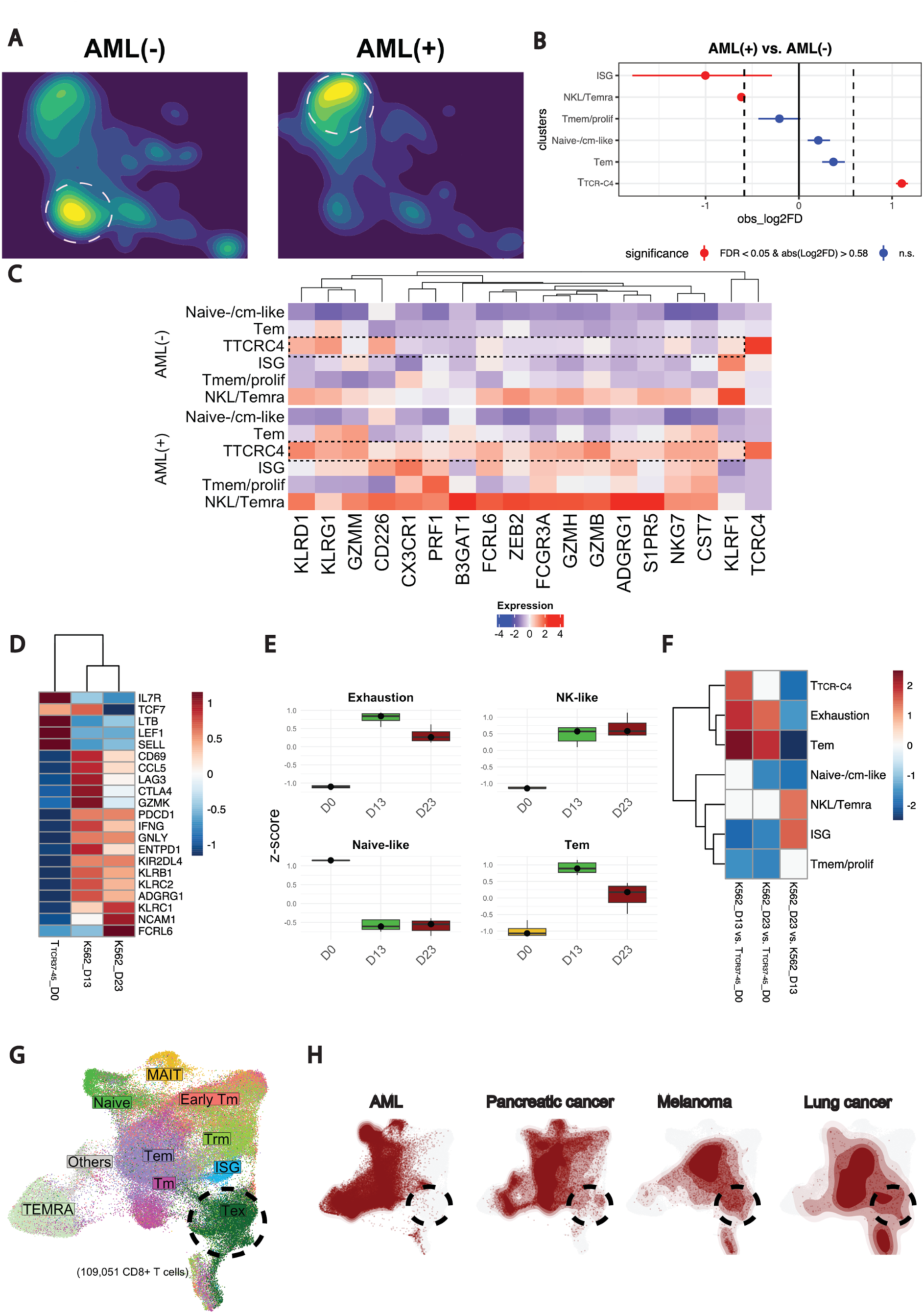
AML induces T_TCRC4_ NKL/Temra differentiation skewing. **(A)** UMAP plot of CD8^+^ T cells colored by density and split by group (AML(-) and AML(+)). AML(+) samples are those in which leukemic cells were detected in BM or PB, while AML(-) samples lack detectable leukemic cells. The plot shows the two-dimensional distribution of CD8+ T cells, with color intensity representing cell density, where blue indicates lower density and yellow indicates higher density. Dashed lines highlight areas of highest cell density in both AML(+) and AML(-) groups. **(B)** Point-range plot showing the pairwise (AML(+) vs. AML(-)) proportional difference for each CD8^+^ T-cell subset. Colors indicate the statistical significance (red: FDR < 0.05, blue: FDR >= 0.05); vertical dashed lines mark the absolute value of log2FD cutoff for significance. **(C)** Heatmap displaying DGE of NKL genes across the CD8^+^ T-cell subsets split horizontally by AML(-) vs. AML(+), with each condition showing the gene expression patterns for CD8+ T-cell subsets. Blue indicates low expression, while red indicates high expression. Dashed boxes highlight NKL/Temra genes, emphasizing their relative expression across the two conditions. **(D)** Heatmap depicting significant DEGs across three conditions: T_TCR37-45__D0 (T_TCR37-45_-only), K562_D13 (T_TCR37-45_ T cells after 13 days of coculture with K562 AML cell line), K563_D23 (T_TCR37-45_ T cells after 23 days of coculture with K562 AML cell line). Signifcance was determined using the Likelihood Ratio Test in DESeq2,^80^ with genes filtered for adjusted p-value (FDR < 0.05). Color intensity represents relative gene expression, with blue indicating low expression and red indicating high expression. **(E)** Boxplots illustrating the z-score of Exhaustion, NK- like, Naïve-like and Tem signatures across the three conditions (co-culture timepoints). **(F)** Heatmap illustrating the enrichment of scRNAseq-derived (T_TCRC4_, Tem, Naïve.like, NKL.Temra, ISG, Tmem.prolif) and manually curated exhaustion markers across three conditions: K562_D13 vs. T_TCR37-45__D0, K562_D23 vs. T_TCR37-45__D0, K562_D23 vs. K562_D13. The signatures shown are derived from the top 50 DEGs for each subset from scRNAseq dataset. Additionally, a manually curated exhaustion signature is included. Each row represents the relative enrichment of these gene signatures in the comparison of interest, while the color intensity indicates the degree of enrichment: red indicates high enrichment, while blue indicates low enrichment. The comparison group correspond to timepoint during coculture with the K562 cell line. T_TCR37-45_ represents the baseline (T_TCR37-45_-only condition) **(G)** UMAP plot showing the distribution of CD8^+^ T-cell states from a published independent dataset used as reference atlas.^35^ Dashed line indicates the UMAP coordinates of Tex CD8^+^ T-cell subset. **(H)** UMAP plot of CD8^+^ T-cell reference atlas (light grey) overlaid with projections (dark red) of CD8^+^ T cells collected from various independent published scRNAseq datasets (queries) of AML^20,22,23,44,45^, pancreatic cancer,^46^, melanoma, ^47^ and lung cancer.^48^ Dashed line indicates the area corresponding to Tex.

Following previously described methodology,^42^ we exposed *in vitro* CD8^+^ transgenic WT1-specific T cells targeting the HLA A*0201-restricted WT137-45 epitope^43^ (T_TCR37-45_) to high-WT1-expressing HLA- A*0201-transduced K562 acutely transformed chronic myelogenous leukemia (**Extended data Fig. 11A**). As K562 primarily express standard proteasomes and are not readily lysed by T_TCR-C4_ targeting the immunoproteasome-specific WT1126–134 peptide, we used T_TCR37-45_ due to its proteasome-agnostic WT1- derived peptide recognition.^43^ Exposing T_TCR37-45_ to fresh K562 every 3-4 days at stable effector-to-target (E:T) ratios (1:1, 1:4) caused initial tumor lysis **(Extended data Fig. 11B)**, but control was lost by day 13, marked by a declining T_TCR37-45_-to-tumor ratio **(Extended data Fig. 11C)**. Irrelevant TCR-expressing T cells exerted no control **(Extended data Fig. 11D)**. Bulk-RNA sequencing on sorted T cells found increased (FDR <0.05) naïve/stem-like genes (*IL7R, TCF7, LTB*, *LEF1*, *SELL*) at day 0, followed at day 13 by increased effector (*CCL5, GZMK*), activation/exhaustion (*CD69*, *LAG3*, *CTLA4*, *PDCD1*), and NKL/Temra markers (*KLRB1*, *ENTPD1*, *KLRC1*, *KLRC2, NCAM1*) (**Fig. 4D-E**). By day 23, the activation/exhaustion markers decreased, while NKL/Temra persisted or increased (**Fig. 4D-E**). To integrate these findings with the scRNAseq dataset, we performed a gene-set enrichment analysis with the top 50 differentially expressed genes from each CD8^+^ subset identified in scRNAseq, including naïve-/cm-like, Tem, NKL/Temra, T_TCR-C4_, Tmem/prolif and ISG from the scRNAseq dataset. We also included a manually curated exhaustion signature **(Supplementary Table 9)**. Exhaustion and Tem signatures exhibited enrichment in later timepoints, yet there was no further increase at day 23 versus day 13 (**Fig. 4F**). In contrast, the NKL/Temra increased between days 13 and 23 corresponding to T_TCR37-45_ loss of tumor control **(Extended data Fig. 11C)**.

We next projected scores from published NKL dysfunction signatures^22,38,39^ onto our UMAP, revealing overlaps that link the NKL/Temra CD8^+^ T cells we identified with an end-term dysfunctional differentiation subset **(Extended data Fig. 12)**. To confirm that AML-exposed T cells skewed towards a NKL/Temra rather than exhausted (Tex) phenotype, we projected onto a scRNAseq reference atlas a BM AML CD8^+^ T-cell dataset, compiled from published sources,^20,22,23,44,45^ alongside datasets representing pancreatic,^46^ melanoma,^47^ and lung^48^ solid tumors (**Fig. 4G-H**).^35^ Unlike solid tumors, which included a Tex cluster, AML datasets aligned with our spectral flow-cytometry and scRNAseq findings confirming the absence of Tex and the presence of NKL/Temra cells (**Fig. 4G**, **Extended data Fig. 13**). Although these studies did not specifically analyze tumor-antigen reactive T cells, the findings collectively suggest that the NKL (versus exhaustion) signature is intricately associated with AML-induced T-cell dysfunction.

### Prolonged azacitidine exposure enhances self-renewal and Tcm features of T_TCR-C4_, supporting TTCR- C4-mediated AML control

Given the link between NKL differentiation and AML-induced T-cell dysfunction, we explored the state of T_TCR-C4_ in patient 8, the only patient of 4 **(Extended data Fig. 4)** who exhibited prolonged T_TCR-C4_ and MRD persistence indicative of disease control despite incomplete leukemia clearance, enabling TTCR- C4/AML interplay analysis. This female patient underwent a non-myelo-ablative (Flu/3Gy-TBI) HCT from a male donor, with MRD (0.26% blasts by flow cytometry) detected 28 days post-HCT (**Fig. 5A**) and received a T_TCR-C4_ (10^10^ cells/m^2^) 77 days post-HCT. AML blasts expressed detectable but limited WT1 protein pre-HCT **(Extended data Fig. 14).** scRNAseq at day 49 post-infusion revealed circulating CD34^+^ female-origin blasts (XIST^+^) expressing WT1 (**Fig. 5E-F**). Persistent MRD and limited T-cell persistence at day 63 post-infusion (T cells declined to only 0.1 multimer^+^ CD8^+^ T cells/µl on day 60), (**Fig. 5B**, **5C- red arrow, 5D)** prompted azacitidine treatment (five days every 28 days). Post-azacitidine, T_TCR-C4_ counts increased (∼50 multimer^+^ CD8^+^ T cells/ul; >3% CD8 multimer^+^ T cells) without additional T_TCR-C4_ (**Fig. 5D**), leading to azacitidine discontinuation after seven cycles when no AML was detected. However, AML recurrence at day 414 post-infusion triggered chemotherapy (mitoxantrone, etoposide and cytarabine), additional azacitidine cycles followed by a second T_TCR-C4_ (10^10^ cells/m^2^) infusion (**Fig. 5A**). Azacitidine was held post-infusion but restarted upon MRD detection 162 days post-second T_TCR-C4_ infusion, continuing for over 20 monthly cycles, maintaining marrow MRD below 1% for 21 months (641 days, from day 868 to day 1509) before progression was detected on day 1546 (**Fig. 5A-B**). During this 21-month period, blasts were detectable via scRNAseq in BM but not in PB (**Fig. 5F**, **Extended data Fig. 15A-B)**. Throughout the azacitidine cycles, platelet counts decreased (∼100 x 10^3^ platelets/µl) mid-cycle before rising (∼300 x 10^3^ platelets/µl) immediately before the next cycle, consistent with transient azacitidine-mediated myelosuppression. Neutrophil counts, however, paradoxically increased mid-cycle (∼4.5 x 10^3^ neutrophils/µl) and decreased (∼1.6 x 10^3^ neutrophils/µl) just before the next cycle, suggesting that treatment-related AML contraction facilitated enhanced mid-cycle neutrophil production despite azacitidine’s myelosuppressive effect **(Extended data Fig. 15C)**. This 21-month response exceeds the typical 4-month median for post-HCT azacitidine treatment,^49^ suggesting that persisting T_TCR-C4_ plus azacitidine contributed to long-term disease control.

**Fig. 5.**
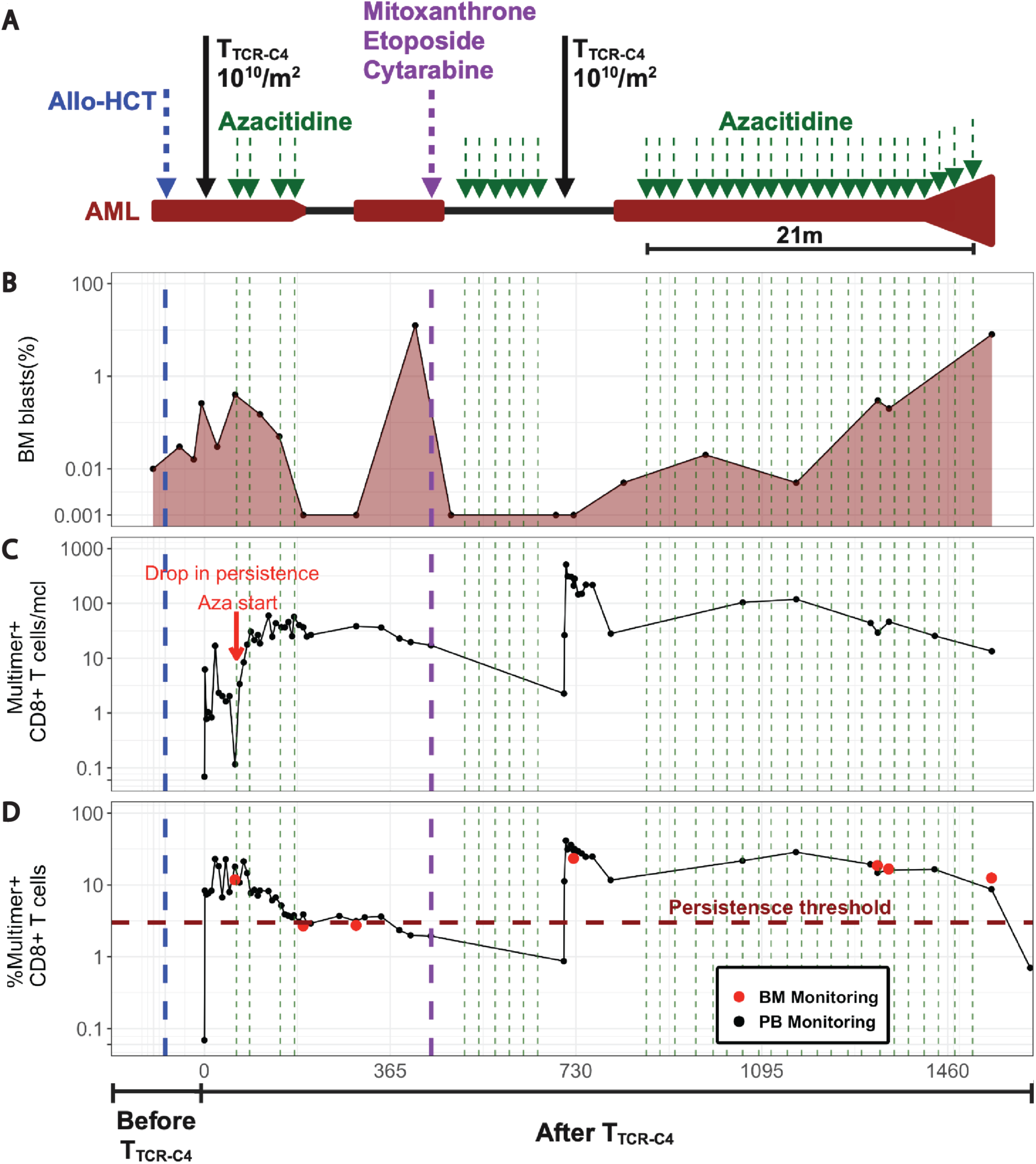
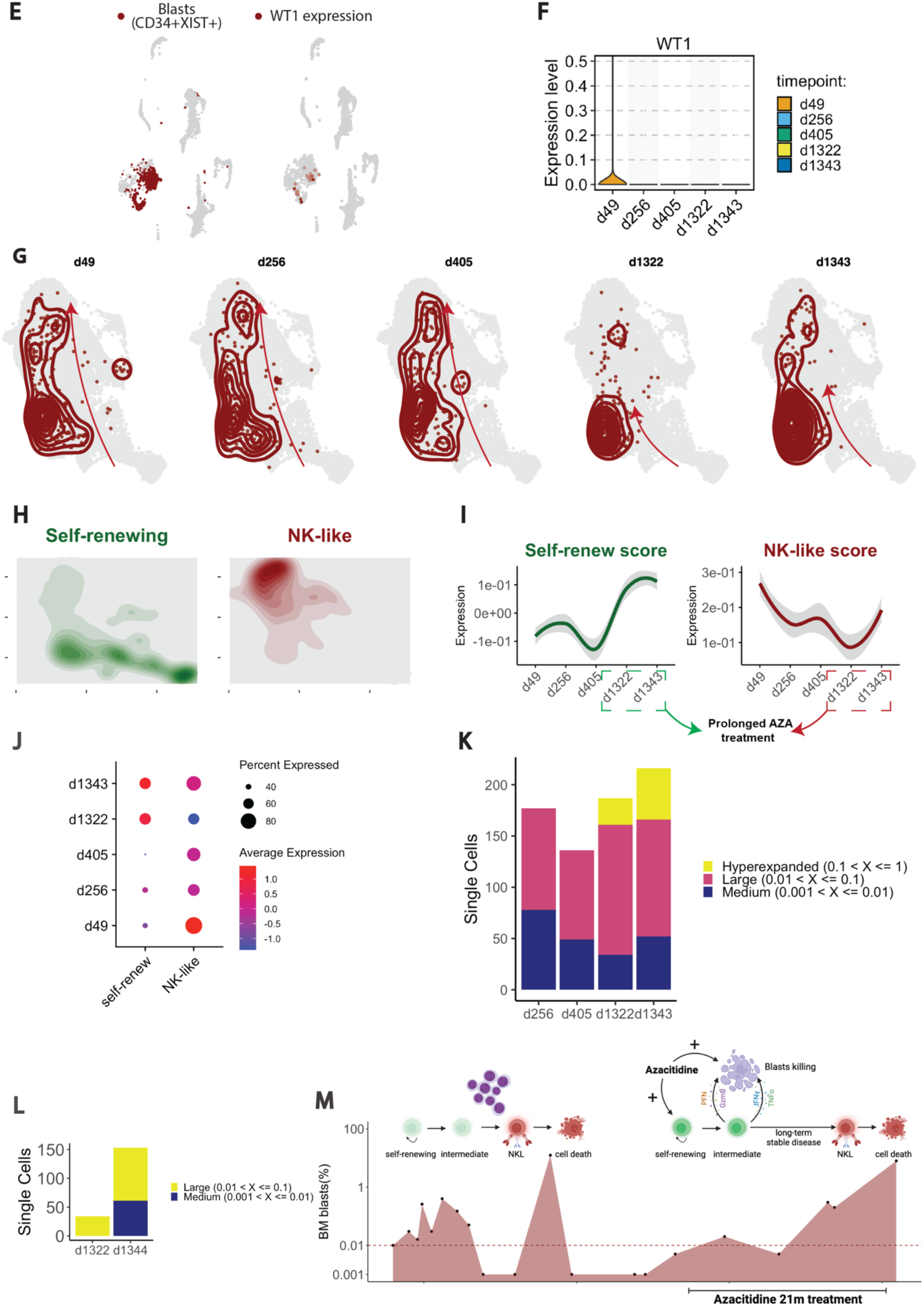
Azacitidine as a potential boost for T_TCR-C4_ *in vivo* function and persistence. **(A)** Timeline of patient’s treatment regimen. Dark red highlights timeframes with detectable BM AML **(B)** Percent of BM AML blasts (y-axis, log scale) by multiparametric flow cytometry (MFC) at specific timepoints (black dots, dark red shaded area). The horizontal dashed line represents the sensitivity limit of MFC. **(C)** Multimer^+^ cells/µl and **(D)** Percent multimer^+^ of CD8^+^ T cells in PB (black dots) and BM (red dots) collected before and after T_TCR-C4_ infusions. The red arrow indicates the lack of T_TCR-C4_ persistence before the start of Azacitidine **(E)** UMAP plots showing a blast score (dark red) calculated based on the co-expression of *CD34* and *XIST* (a female-specific gene) in a patient previously transplanted with a sex-mismatched donor (female patient, male donor). The score was used to identify cells expressing a high blast score. Additionally, *WT1* expression across all cell populations is shown. Cells with high expression of each marker are highlighted in red, while other cells appear in grey. **(F)** Violin plot of the *WT1* expression (y-axis) across the timepoints analyzed after first infusion of T_TCR-C4_ (x-axis, d265, d405, d1322, d1343). Violin plots are colored by timepoint. **(G)** UMAP plots display the distribution of total PB CD8^+^ T cells from the scRNAseq dataset, with the T_TCR-C4_ subset from patient 8 highlighted in dark red. Single-cell points represent individual cells, while kernel density contours (also in dark red) depict the density of T_TCR-C4_ cells within the CD8^+^ T cell landscape. The plots are stratified by timepoints post-infusion: (day 49, day 256, day 405, day 1322, and day 1343). This series illustrates the spatiotemporal dynamics of the T_TCR-C4_ subset, showing its distribution in relation to the broader PB CD8^+^ T cell population and highlighting potential transcriptional changes over time. Red arrows were added manually to indicate the different skewing of T_TCR-C4_ across the timepoints examined. **(H)** UMAP plots showing the distribution of PB CD8^+^ T cells, with the density contours highlighting cells with high self-renewing (dark green) and NK-like (dark red) scores. The self- renewing score was calculated based on a set of stem-like/survival genes (*TCF7*, *LEF1*, *SELL*, *CCR7*, *BCL2*, *IL7R*, *CD27*, *CD28*) and the NK-like score was based on NK-associated genes (*ZEB2*, *S1PR5*, *CX3CR1*, *KLRG1*, *NKG7*, *FCRL6*, *KLRD1*, *ADGRG1*). The plots are color-coded by score (light grey for the background, darker shades indicating higher scores). Cells with scores above the 75th percentile for each gene set are highlighted with density in dark green for high self-renewing score cells and dark red for high NK-like score cells. **(I)** The plot shows the temporal changes in the self-renew (left) and NK-like (right) scores of T_TCR-C4_ from patient 8. The smoothed trends of the scores were calculated using LOESS regression at multiple timepoints. The dark green (self-renew) and dark red (NK-like) lines represent the smoothed score trends, with shaded areas indicating the confidence interval. The y-axis reflects the score expression values. **(J)** Dot plot displaying the expression of the self-renew and NK-like scores across timepoints post-infusion. The size of each dot corresponds to the expression level of the respective score and the color indicates the relative intensity of the expression. **(K)** Stacked plot showing cell count of T_TCRC4_ in specific clonal frequency ranges over time (d256, d405, d1322, d1343). Colors indicate the clonal frequencies. **(L)** Stacked plot showing cell count for BM T_TCRC4_ within specific clonal frequency ranges at d1322 and d1343. Colors indicate the clonal frequencies. **(M)** Cartoon (created with Biorender) showing the influence of blasts and azacitidine on T_TCR-C4_. The red area under the curve represents the blasts percentage (y-axis) over time (x-axis). Blasts induce T_TCR-C4_ skewing towards NKL and cell death. However, prolonged exposure to azacitidine allows T_TCR-C4_ to maintain self-renewal, facilitating persistence and long-term disease control.

T_TCR-C4_ transcriptomics were analyzed to identify associations with disease control. We scored genes associated with self-renewal and NK-like profiles and found that at earlier timepoints (d49, d256, and d405 post-first infusion), T_TCR-C4_ were predominantly skewed towards an NKL/Temra transcriptional state. However, at later timepoints (d1322, d1343) although T_TCR-C4_ remained along the same differentiation trajectory (from less differentiated states to NKL/Temra T_TCR-C4_), they exhibited fewer NKL features, and showed an enrichment of self-renewal markers (**Fig. 5G-I**). scTCRseq analysis of PB at days 265, 405, 1322, 1343 post-first T_TCR-C4_ infusion revealed progressive clonal expansion of T_TCR-C4_ (**Fig. 5K**). T_TCR-C4_ clonal expansion peaked during the 21 months of continued azacitidine treatment, suggesting that during this phase, T_TCR-C4_ were capable of both self-renewal, which in turn facilitated their long-term persistence, and antigen recognition, which triggered further expansion and disease control. Similarly, BM scRNAseq revealed clonal expansion of T_TCR-C4_ between d1322 and d1344 post-first infusion (**Fig. 5L**) despite no additional T_TCR-C4_. These findings indicate that azacitidine-exposed T_TCR-C4_, retained a self-renewing (versus NKL) features, which enabled their long-term persistence and enhanced AML-targeting efficacy that may have contributed to long-term disease control (**Fig. 5M**).

## Discussion

We previously demonstrated that EBV-specific T_TCR-C4_ infusion prevented AML recurrence in patients at high risk of post-HCT relapse.^9^ In 15 AML patients who received T_TCR-C4_ post-HCT relapse, we observed indirect anti-leukemic activity, but no survival advantage in those receiving EBV- or CMV- specific T_TCR-C4_. We analyzed the AML/T_TCR-C4_ interplay to investigate how AML affects antigen-specific T cells, and identified T-cell characteristics associated with AML control in one case.

The characteristics of substrate cells from which T-cell products are derived can impact post-transfer persistence. In murine and macaque models, antigen-specific Tcm-like CD8^+^ T cells persist long-term post-adoptive transfer, unlike Temra-like cells.^9,28^ In our study, T_TCR-C4_ from EBV-specific persisted longer compared to CMV-specific substrate cells which may reflect the distinct biology of EBV and CMV.^50,51^ CMV reactivates periodically, requiring rapidly activated responses that promote Temra differentiation,^52,53^ while EBV tends to remain quiescent, preserving Tcm phenotypes.^54–56^ While reactivation frequency plays a role, the reactivation microenvironment is also critical. CMV reactivates in non-professional antigen-presenting cells, including fibroblasts, which lack co-stimulatory signals and favor Temra differentiation; EBV reactivates in lymphoid tissues B cells, providing co-stimulation that may preserve the Tcm phenotype.^57,58^ Our analysis of an independent mass-cytometry dataset^29^ confirmed that EBV-specific cells better maintain self-renewing features than CMV-specific cells. Taken together these findings establish that self-renewal may predict prolonged T-cell persistence.^9,28^ However, EBV-specific T_TCR-C4_ were unable to sustain long-term responses in all patients, suggesting influence by non-intrinsic AML-related factors.

Despite efforts to investigate T cell dysfunction in AML,^20,21,23^ several critical questions have remained unanswered,^25^ mainly due to the challenge of identifing AML-specific T cells. In this study, infused T_TCR-C4_ offered the possibility of examining AML-specific T cell and AML biology. We found that T-cell exhaustion is not the primary cause of AML-specific T-cell dysfunction.^20,21,23^ Instead, T_TCR-C4_ with exhaustion markers (PD1, TIGIT, Tim3) also expressed activation markers (CD38, CD69) and proliferation (Ki67), indicating an effector-like T-cell state. Overall, this suggests that AML-exposed T cells follow a differentiation trajectory from effector-memory to NKL cells, compromising T_TCR-C4_ cell persistence and function.

AML’s distinct properties, including myeloid cells immunosuppressive effects^59^ via production of reactive oxygen species and other soluble factors, may distinctly redirect T cells toward functional impairment that may preclude transitioning to a classical exhausted phenotype and explain the observed NKL skewing in T_TCR-C4_ and endogenous CD8^+^ T cells. The absence of classical exhausted cells in AML has several implications. In solid tumors, chronic antigen stimulation induces a multi-step epigenetic shift of CD8^+^ T cells into exhaustion,^60^ starting from PD1^+^TCF1^+^ precursor/progenitor exhausted cells,^23,24^ whose frequency correlates with response to checkpoint inhibitors.^9,28^ In contrast, our AML findings suggest that exhaustion markers reflect recent T cell activation rather than true exhaustion, which may also explain the limited efficacy of checkpoint inhibitors in AML.^20,23,24,61^ The presence of a distinct dysfunctional program supports new immunotherapy strategies to enhance the efficacy of anti-AML adoptive T-cell therapy, preventing T-cell dysfunction. For example, knocking out ID3 and SOX4 transcription factors in chimeric antigen receptor-engineered (CAR)-T cells *in vitro* reduces NKL skewing and enhances effector functions.^42^ Whether these strategies can improve cell therapy in AML *in vivo* remains to be determined.

Another strategy to reduce NKL skewing of T_TCR-C4_, as suggested by the outcome in one of our treated patients, could involve the hypomethylating agent azacitidine. While azacitidine has direct anti-leukemia effects,^62^ the prolonged disease control observed in our patient was unusual prompting further investigation into the mechanisms at play. Azacitidine administration correlated with clonal expansion and persistence of central memory, as opposed to NKL differentiated T_TCR-C4_, leading to a prolonged equilibrium between anti-leukemic T_TCR-C4_ and AML MRD. Before the first T cell infusion in this patient, low AML WT1 expression likely did not activate T_TCR-C4_, causing their near disappearance. However, after azacitidine introduction, T_TCR-C4_ frequencies increased, likely driven by azacitidine-induced WT1 expression.^63^ Intermittent T_TCR-C4_ activation by WT1 presentation on AML cells transiently induced by cycles of azacitidine with subsequent temporary clearance of AML may have helped sustain T_TCR-C4_ persistence and response to stimulation. Other studies have shown that azacitidine can have negative effect on T regulatory cells^64^ and boosts CAR T cell toxicity towards AML.^65^ Further investigation is needed to determine if azacitidine acts preferentially on leukemia cells by increasing WT1 expression thereby promoting antigen-recognition and/or T_TCR-C4_ effector functions. However, taken together, these findings support azacitidine as a favorable adjunct to T cell immunotherapy.

Paradoxically, our ability to track dysfunction was limited to patients with persisting T_TCR-C4_, where T cells were relatively more functional. We hypothesize that patients with more aggressive disease experience accelerated T-cell dysfunction and death rather than establishment of progenitor cells capable of self-renewal; however we were unable to formally test this in our study. Despite a limited sample size, our findings on the nature of T-cell dysfunction in AML were confirmed in larger cohorts of endogenous CD8^+^ T cells, though further validation in larger trials could provide more detail. Additionally, our interpretation of the effect of azacitidine on T_TCR-C4_ is based on a single patient. To address this limitation, we are initiating a clinical trial of co-administration of azacitidine and TCR-T cells.

Our study illuminates some of the complex mechanisms underlying AML-induced T cell dysfunction and strengthens support for a distinct pathway outside the traditional paradigm of T cell exhaustion, emphasizing the need to addres this unique dysfunction in future immunotherapy strategies.

## Methods

### Clinical protocol

The trial was approved by the Fred Hutchinson Cancer Center (FHCC) Institutional Review Board, the US Food and Drug Administration and the National Institutes of Health Recombinant DNA Advisory Committee. It was registered at ClinicalTrials.org under the identifier NCT01640301. Eligible participants included ‘high-risk’ AML patients with relapsed or refractory disease (overt or MRD) post-HCT along with their fully HLA-matched (10 of 10) related or unrelated donors expressing HLA A*0201 (HLA-A2).

### Patient selection

HLA-A2 genotype was confirmed by high-resolution typing before enrollment. Exclusion criteria included: refractory central nervous system disease, HIV seropositivity, grade ≥ 3 GVHD and no available CMV/EBV-seropositive matched donor. The sample size for this study was not based on formal power calculations, but on feasibility, the potential to provide descriptive information, determine whether further study was warranted and evaluate toxicity.

### Treatment plan

Isolation of TCRC4, lentiviral vector construction, and T_TCR-C4_ generation were performed as previously described.^9^ Patients were eligible to receive a first infusion of T_TCR-C4_ after demonstrating relapse (overt or MRD) at any time post allogeneic HCT. Patients 26,27,28 received lymphodepleting treatment before the first T_TCR-C4_ infusion with cyclophosphamide (300 mg/m^2^ IV) and fludarabine phosphate (30 mg/m^2^ IV) daily on days -4 to -2. A second T_TCR-C4_ infusion was administered only if the frequency of T_TCR-C4_ was <3% of total peripheral CD8^+^ T cells. Patients 5,7,19,23,27 received 1 infusion, patients 4,8,15,26,28 received 2 infusions, patient 1 received 1 infusion, the remainder received 4 infusions (**Table 1**). Patients were monitored for toxicities, based on Common Toxicity Criteria v.4.0.

### Assessment of disease status

Morphology, multiparameter flow cytometry, standard cytogenetics or genomic technologies were routinely performed on bone marrow aspirates and peripheral blood samples that were obtained from all patients. Any level of residual disease was considered to indicate positivity for MRD.

### T cell tracking by WT1 peptide/HLA (pHLA) tetramers

WT1 pHLA-specific tetramers (produced by the FHCC Immune Monitoring Core Facility) were used to detect T_TCR-C4_ in PBMCs collected after infusions, with a staining sensitivity of 0.01% of total CD8^+^ T cells, as previously described.^9^ T_TCR-C4_ percentages were calculated using FlowJo v.10 (Treestar).

### Patient outcomes and survival analysis

Kaplan-Meier OS curves were estimated using the *survminer* (v0.4.9) and *survival* (v3.3-1) packages in R. OS was calculated from the date of first T_TCR-C4_ infusion to the date of death or censoring. Outcomes of responding patients were represented using a swimmerplot. Day 0 was defined as the post-HCT relapse date or, for HCT-refractory patients, as day 28 post-HCT, at which timepoint persistent disease was observed. R package *swimplot* (v1.2.0) was used for visualization.

### Analysis of T_TCR-C4_ persistence

Persistence of TCR-T cells (%T_TCR-C4_) over time was visualized using *ggplot2* (v3.4.4) R package. Wilcoxon rank-sum tests were used for pairwise comparisons between T_TCR-C4_ with different virus-specificity (EBV vs. CMV) over time. Kruskal-Wallis tests were applied for multi-group comparisons.

Patients were categorized based on disease status prior to infusion: those with detectable MRD or overt disease before T_TCR-C4_ infusion, or those who relapsed within 3 months post-HCT, were classified as having "aggressive" disease. Patients without these risk factors were considered "non-aggressive." One patient was excluded from the analysis due to the unavailability of WT1-expression data, which could confound interpretation of the persistence results. Fisher’s exact test was used to evaluate the association between disease aggressiveness and T_TCR-C4_ persistence. The test was chosen due to the small sample size and binary classification of persistence (long-term vs. short-term) and disease aggressiveness (aggressive vs. non-aggressive). Statistical significance was assessed with a p-value threshold of 0.05.

### Flow-cytometry

Cryopreserved PBMCs were thawed and allowed to rest overnight in RPMI medium supplemented with 10% fetal bovine serum (R10). The cells underwent stimulation with a cocktail containing the WT1126-134 peptide at a final concentration of 1 μg/ml in R10 and intracellular cytokine staining using a 15-color staining panel, as previously described.^9^ Flow cytometry was conducted on an LSRII instrument (Becton Dickinson) with data acquisition using FACS-Diva software v.8.0.1. Flow cytometry data were subsequently analyzed using FlowJo v.10 (Treestar). Finally, the percentage of Tetramer^+^cytokine^+^ cells gated on the CD8^+^ T cell population was determined.

### Spectral flow cytometry

Post-infusion PB samples were analyzed using a 5-laser Cytek Aurora. Antibodies used are listed in **Supplementary Table 1**. T_TCR-C4_ were identified by binding to the APC dye-labeled HLA-A2:WT1126-134 tetramer. Spectral flow-cytometry data were biexponentially transformed, compensated and preprocessed (aggregates and dead cell removal) in FlowJo V10 (TreeStar). Pregated CD8^+^ T cells were exported from FlowJo and loaded in R (v4.3.2). First we created a flowSet using *flowCore* (v2.12.2)*^66^*and subsequently analyzed the data using *CATALYST* (v1.24.0).^67^

Ki67 was then used to manually gate subsets of interest using the FlowJo software. Statistical analysis included Kruskal-Wallis to compare T_TCR-C4__Temra and T_TCR-C4__Tem percentages over time, and Wilcoxon test for comparing KLRG1^+^CD57^+^GNLY^+^ and Tim3^+^PD1^+^TIGIT^+^ T_TCR-C4__Temra.

### *In-silico* mass cytometry validation

Mass cytometry files used to generate an atlas of virus-specific CD8^+^ T cells^29^ were downloaded from https://zenodo.org/records/8330231. Data were analyzed using *flowCore* and *CATALYST* R packages as specified above.

### Single Cell RNA Sequencing

Patients with persisting PB T_TCR-C4_ (> 3% of the total CD8^+^ T cells) at least until day 28 after infusion were selected for scRNAseq analysis. Available PBMCs or BMMCs were thawed, washed and loaded on a 10x Chromium Controller based on the 3’ Chromium or 5’ Chromium Single Cell V(D)J Reagent Kit manual (10x Genomics). Library preparation was performed as per manufacturer’s protocol with no modifications. Library quality was confirmed by TapeStation High Sensitivity (Agilent, evaluates library size), Qubit (Thermo Fisher, evaluates dsDNA quantity), and KAPA qPCR analysis (KAPA Biosystems, evaluates quantity of amplifiable transcript). Samples were mixed in equimolar fashion and sequenced on an Illumina HiSeq 2500 rapid run mode according to the standard 10X Genomics protocol. TCR target enrichment, 5’ gene expression library, and TCR library were carried out to the 5’ Chromium Single Cell V(D)J Reagent Kit manual (10x Genomics). The 10X Genomics software Cell Ranger (v2.0.0) was used to process the raw data FASTAQ files with default parameters. The EmptyDrops method^68^ was used to identify cells with low RNA contents. The “count” function was used to perform alignment, filtering, barcode counting and UMI counting. Reads were aligned to the hg38 human reference genome (Ensembl) and the known transgene codon-optimized sequence using Spliced Transcripts Alignment to a Reference (STAR).^69^

For V(D)J sequencing assembly and paired clonotype calling, we used CellRanger “vdj” function. This function leverages Chromium cellular barcodes and UMIs to assemble V(D)J transcripts for each cell. CellRanger V(D)J calling produces an output named “filtered_contig_annotations.csv” for each sample, which lists CDR3 amino acid and nucleotide sequences for single cells identified by their barcodes.

### scRNAseq quality control and subsetting

The filtered feature matrices generated by the CellRanger pipeline were used for downstream quality control (QC) and analyses. We used the function read10xCounts from the R package *DropletUtils* (version 1.14.2) to load the CellRanger output in R as a SingleCellExperiment^70^ object. Doublet cells filtering was performed on each sample using the *scds* package (v1.10.0).^71^ QC and filtering were conducted using the *scater* R package (v1.22.0)^72^.^71^ Genes not detected across all the cells were removed and cells were filtered based on feature counts, the percentage of mitochondrial and ribosomal genes, and the number of expressed features. Cells with values beyond a specific threshold, between 1 and 3.5 median absolute deviations (MAD) from the median, were excluded. These MAD thresholds were established according to the quality of each sample. Features with a count greater than 1 in at least 3 cells were retained for downstream analysis. We then split cells by sample in 15 datasets, normalized, found the 2000 most variable genes and scaled for each dataset using *Seurat* (v4.3.0.9001) SplitObject, NormalizeData, FindVariableFeatures and ScaleData respectively.^73,74^ For batch correction, we used the FindIntegrationAnchors and IntegrateData functions from *Seurat*. The integrated dataset was then used for scaling (ScaleData) and dimensionality reduction (PCA and UMAP) using RunPCA and RunUMAP, respectively. UMAP dimension reduction and clustering were computed using the first 20 principal components (PCs). The number of PCs capturing most of the variation in our data was selected using *Seurat* function ElbowPlot which visualizes the standard deviation of each PC. Clusters were identified via shared-nearest-neighbor-based (SNN) clustering and further analyzed at a resolution of 0.6. CD8^+^ cell subset was identified using *scGate* (v1.0.1)^34^ and subsequently extracted with Seurat subset function.

### scRNAseq CD8^+^ endogenous and T_TCR-C4_ analysis

The CD8^+^ subset was used to find the 2000 most variable features (FindVariableFeatures), scale the data (ScaleData), and run dimensionality reduction (runPCA and runUMAP). SNN was used for clustering with a resolution of 0.5 for downstream analysis. Differential gene expression across clusters or condition (e.g. AML(+) vs AML(-)) was computed using *Seurat* FindAllMarkers with default parameters. TCRC4- transgene^+^ cells were identified using *scGate* (v1.0.1). For visualization purposes, we used Seurat built-in functions alongside *ComplexHeatmap* (v2.15.1)^75^, *scCustomize* (v1.1.0.9001) (https://github.com/samuel-marsh/scCustomize) and *SCP* (https://github.com/zhanghao-njmu/SCP). The pan-cancer CD8^+^ single-cell reference atlas was built using the CD8.thisStudy_10X.seu.rds file^35^ downloaded from Zenodo and processed using the make.reference function from the *ProjecTILs* R package (v3.2.0).^76^ We then used Run.Projectils function, from the same package, to project the cell states from the query dataset onto the reference.

Cell trajectory inference was computed using the *Monocle* R package (v1.3.4), with UMAP used as dimensionality reduction. Single-gene expression patterns along the pseudotime were visualized using the plot_genes_in_pseudotime function in *Monocle*. RNA velocity was conducted by exporting the CD8^+^ T- cell Seurat object as an h5ad file using the R package *seurat-disk* (v0.0.0.9020) (https://github.com/mojaveazure/seurat-disk), loading this file in Python (v3.8.14) as AnnData object,^77^ and estimating velocities with the *scvelo* Python package (v0.2.4)^41^ using the deterministic model.^77^

We classified patients as AML(+) or AML(-) based on the presence of detectable blasts in BM or PB. Three patients lacked BM or PB evaluations at one timepoint each. For two of these patients, we used the donor-recipient (male-female) sex mismatch. Patient 8 at day 49 exhibited cells expressing the female-specific gene *XIST* alongside the AML-associated genes *CD34* and *WT1* **(Extended data Fig. 15A)**; similarly, patient 25 expressed *XIST* in a *RPS4Y1* (male-specific gene) negative region at days 7 and 28, thus classified as AML(+) at these timepoints **(Extended data Fig. 15B)**. Patient 4 was categorized as AML(-) at day 100 and AML(+) at day 581, as previously reported.^43^

To assess the significance of differences in cell proportions per CD8^+^ cell state between groups (AML(+) vs AML(-)), a permutation test was applied. Specifically, the permutation_test and permutation_plot functions from the R package *scProportionTest* (version 0.0.0.9000) (https://github.com/rpolicastro/scProportionTest) were used with default parameters.

For scRNAseq *in-silico* validation, we compiled scRNAseq datasets from independent studies on AML^20,22,23,44,45^, lung cancer^48^, pancreatic cancer^46^ and melanoma.^47^ CD8^+^ T cells were identified and extracted using the *scGate*^34^ R package. These cells were then projected onto the pan-cancer CD8^+^ T cell reference atlas^35^ using *ProjecTILs* R package.

The TCR repertoire was analyzed using the R package scRepertoire (v1.10.1).^78^ Initially the degree of single cells clonal expansion was defined based on the number of cells sharing the same clonotype: Hyperexpanded (100 < X <= 500), Large (20 < X <= 100), Medium (5 < X <= 20), Small (1 < X <= 5), Single (0 < X <= 1) The function occupiedscRepertoire was used to visualize the degree of clonal expansion by cell-state over time.

### In-vitro chronic antigen stimulation model

The in-vitro dysfunction model was established as previously described.^42^ The WT1^+^ K562 tumor cell line was transduced with lentiviral constructs to express HLA-A*02:01 and GFP, then sorted for purity using the Sony MA900 cell sorter. Cells where then cultured in media consisting of IMDM with GlutaMAX (Gibco, Life Technologies, #31980030) with 10% FBS, and 1% of penicillin/streptomycin.

T_TCR37-45_ cells were obtained following previously published protocols.^43^ We selected TCR-T cells targeting WT137-45 as K562 cell line primarily express the standard proteasome and is not lysed by T_TCR-C4_ targeting WT1126-134.^43^ T cells expressing an irrelevant virus-specific TCR were used as negative control. Co-cultures of T cells with K562 cells were established at a 1:1 and 1:4 E:T ratio (2.5 x 10^5^ T cells, 2.5 x 10^5^ or 1 x 10^6^ tumor cells, respectively). After 3-4 days of coculture, 250 μl of the cell suspension was used for T cell counting and flow-cytometry staining. The remaining cell suspension was spun down, and cells were resuspended in fresh media. A Novocyte 3 lasers flow cytometer was used to quantify GFP^+^ tumor cells and T cells and to maintain constant E:T ratios by reseeding K562 cells. Notably, during the peak of T-cell expansion (day 9-13), the volumes of the cocultures were reduced to ensure the reseeding of an adequate number of tumor cells, thereby maintaining constant E:T ratios. This protocol was followed for 23 days.

### Bulk RNA sequencing

Bulk RNA sequencing was performed using BGISEQ-500 platform at BGI Genomics. Briefly, total RNA was extracted using the Qiagen RNeasy Micro Kit according to the manufacturer’s instructions. For the construction of low input polyA mRNA-seq libraries, the *SMARTseq* (v4) Package was used. Sequencing was performed on a DNBseq T7 machine (MGI) with paired-end 150 bp reads, generating 30M raw reads per sample. Raw sequencing data were filtered and trimmed using the software Soapnuke developed by BGI Genomics. The filtered reads were then aligned to the reference transcriptome using *Bowtie2* (v2.2.5). Gene read counts were subsequently generated from the alignment results using *RSEM* (v1.2.8).

To assess differences over time across conditions (T_TCR37-45__D0, K562_D14, and K562_D23), we used the likelihood ratio test (LRT) with a full model of ∼Condition and a reduced model including only the intercept (reduced = ∼ 1). Normalized counts were obtained using the rlog transformation, and differential expression analysis was performed using the R package *DESeq2* (v1.42.0). We manually curated a list of genes of interest (**Supplementary Table 10**) and filtered the results to retain only those genes that were significant (padj < 0.05) with an absolute log2 fold-change > 1). For visualization purposes, average normalized counts of biological replicates were calculated using the avereps function from the R package *limma* (v3.56.2) and visualized using the R package *pheatmap* (v1.0.12). To visualize the expression patterns of manually curated gene signatures, we calculated z-scores for the averaged normalized counts across conditions (T_TCR37-45__D0, K562_D14, and K562_D23). Finally, to compare the enrichment of scRNAseq-derived gene signatures (top 50 differentially expressed genes) and of a manually curated exhaustion signature across conditions, we used the *hciR* (v1.7) function fgsea_all with default parameters and plot_fgsea for plotting.

## Data sharing

For original data, please contact. Raw data will be deposited and made publicly available as per journal requirements.

## Data Availability

Raw data will be deposited and made publicly available after peer-reviewed publication

## Acknowledgements

We thank all members of the Chapuis and Greenberg lab for their contribution to the manuscript; the Fred Hutchinson Cancer Center Good Manufacturing Practice Cell Processing Facility for generating T_TCR-C4_; the Immune Monitoring Laboratory for generating tetramers; the Flow Cytometry Facility for providing instruments and assistance in flow cytometry assays. We received funding from grant no. P01CA18029-41 (P.D.G.), grant no. NIH-5K08CA169485 (A.G.C.), NIH-T32CA080416 (M.C.L.), the Immunotherapy Integrated Research Center at the Fred Hutchinson Cancer Center (A.G.C., F.M.), Damon Runyon (A.G.C.), the Guillot Family ZachAttacksLeukemia Foundation, Parker Institute for Cancer Immunotherapy (P.D.G.), Gabrielle’s Angel Foundation, the V Foundation, and Juno Therapeutics. We thank all the patients who participated in this study.

## Contributions

Conception and design were performed by P.D.G., A.G.C., F.M., M.B., and T.M.S. Collection and assembly of data were carried out by F.M., L.M., A.G.C., D.N.E., M.B., V.V., D.H. Data analysis and interpretation were performed by F.M., L.M., T.T., V.V., R.G., F.M., A.G.C., and P.D.G. All of the authors contributed to the writing of the manuscript. Final approval of the manuscript was given by all of the authors.

---

**Extended data Fig. 1:**
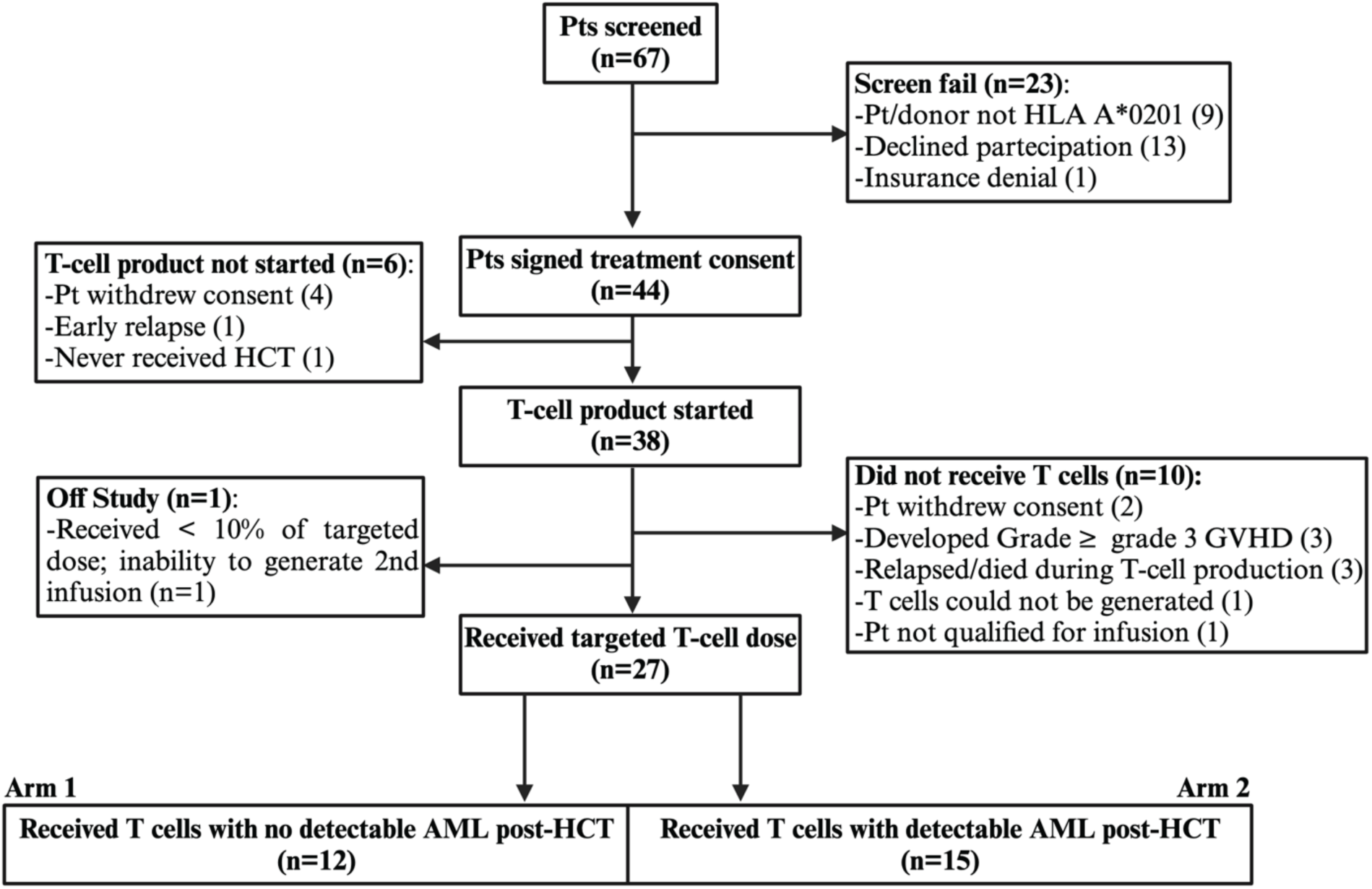
Flow diagram of patients enrolled on the clinical study. Follow-up of the 67 patients screened for participation in the study and the 44 patients who signed the informed consent. Patients with no detectable disease post-HCT were assigned to the Prophylactic Arm^1^, while those with evidence of disease post-HCT were treated in the Treatment Arm described here. A total of 44 pt/donor pairs were enrolled pre- HCT, TCRC4 transduced cells were initiated for 38 patients, among whom 12 disease-free (Arm 1) and 15 relapsed/refractory (Arm 2) patients received TCRC4 transduced cells. An additional patient was considered not evaluable for response outcomes as this patient received <10 % of the targeted dose and was unable to receive further infusions due to inability to generate additional T cells from the donor.

**Extended data Fig. 2:**
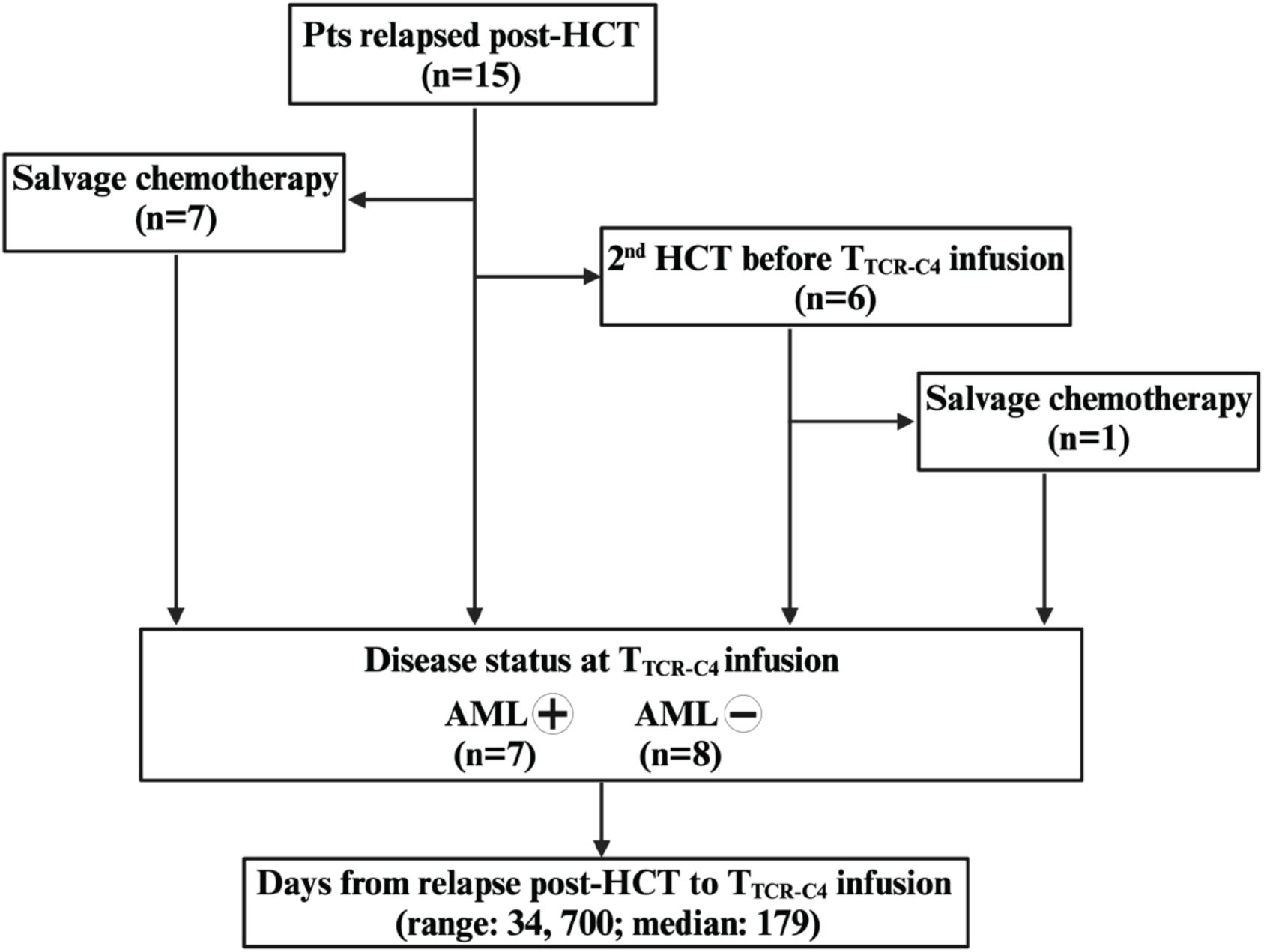
Flow diagram of patients’ treatment history. The cohort included 15 patients who relapsed after HCT. 7 patients received salvage chemotherapy before the infusion of T_TCR-C4_, while 6 patients had two HCTs before the infusion, and 1 patient among them also received salvage chemotherapy. At the time of TCR-C4 infusion, 7 patients had detectable AML, while 8 patients were in remission (AML-negative). The time from relapse post-HCT to TCR-C4 infusion ranged from 34 to 700 days, with a median of 179 days.

**Extended data Fig. 3.**
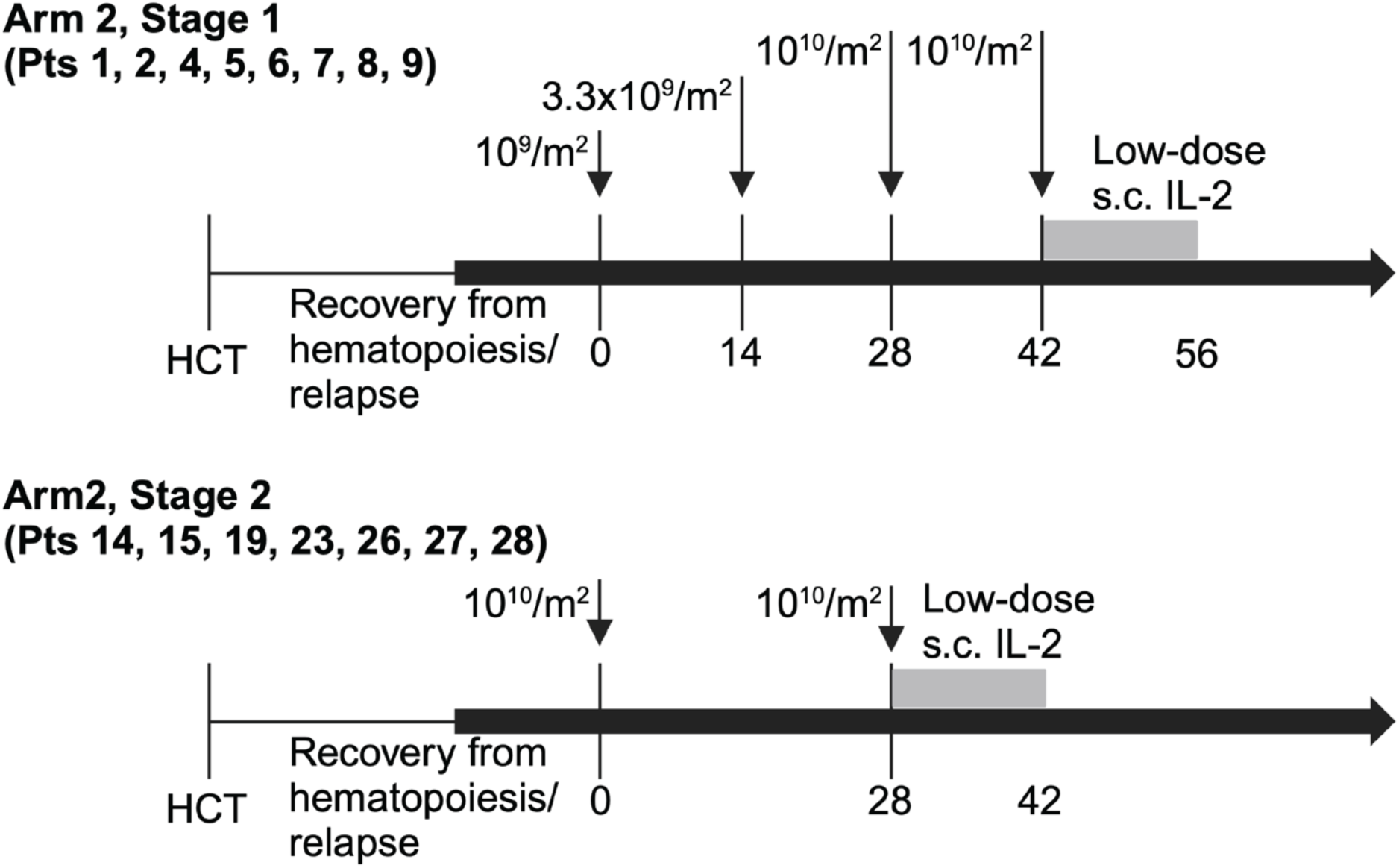
Study schema. The first 8 patients who received T_TCR-C4_ after post-HCT relapse (Treatment Arm, Stage1) received 4 escalating infusions of 10^9^-10^10^/m^2^ 14 days apart, the last infusion followed by s.c. low-dose IL2 for 14 days. After safety of the highest dose was established, all subsequent patients received 2 infusions of 10^10^/m^2^ T_TCR-C4_ 28 days apart, the second infusion followed by s.c. low-dose IL2 for 14 days.

**Extended data Fig. 4.**
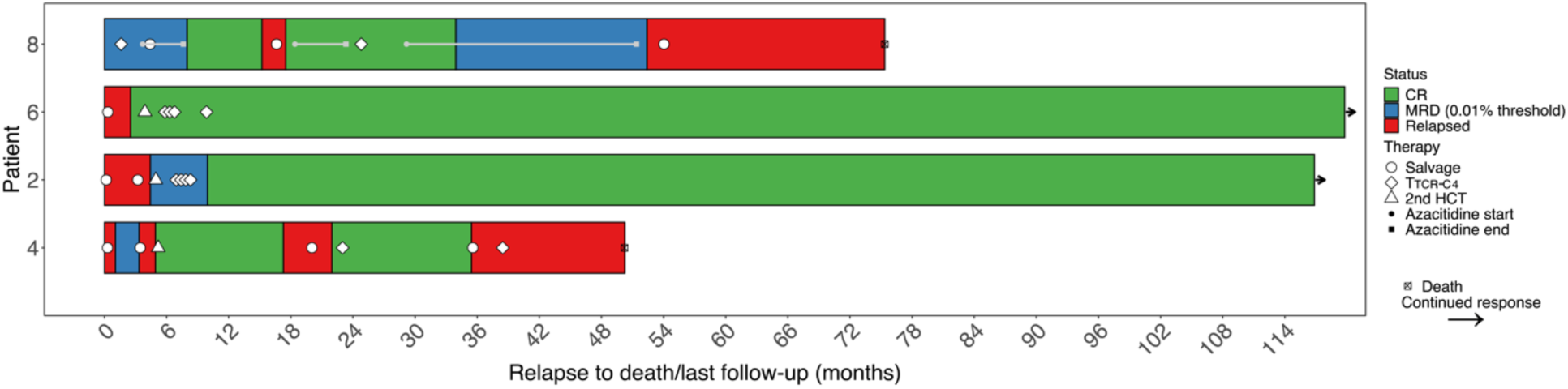
Swimmer Plot of Patient Responses. The swimmer plot illustrates the response duration for patients enrolled in the trial. Time 0 represents the relapse or persistent disease after the first HCT that qualified patients for trial inclusion. The end of each bar indicates the date of death or last follow-up, with arrows marking ongoing responses. "Salvage" denotes any salvage strategy, including chemotherapy, radiotherapy, or intrathecal treatment. Salvage with a second HCT is marked by a triangle. **Patient 8:** A patient who had MRD post HCT and received T_TCR-C4_ with MRD. She later underwent azacitidine salvage treatment for persistent MRD (duration shown as a gray line). The patient then relapsed, underwent additional salvage chemotherapy, azacitidine, and received a second T_TCR-C4_ infusion. Following this, she experienced prolonged MRD+ stable disease with detectable long-term persisting PB T_TCR-C4_. Eventually, the disease progressed, and she relapsed and died (see **Results** for more details). **Patient 6:** A patient who received salvage chemotherapy followed by a second HCT which resulted in no evaluable disease. Subsequently, she received four T_TCR-C4_ infusions. The patient was alive at the last follow-up. **Patient 2:** A patient who, after salvage treatment, underwent a second HCT while in overt disease. She remained MRD-positive 28 days post- second-HCT. She received four T_TCR-C4_ infusions. Eventually, she achieved a MRD negative status. The patient was alive at the last follow-up. **Patient 4:** A patient who achieved a complete remission after a second HCT. However, after ∼15 months, he experienced extramedullary relapse, treated with azacitidine and radiotherapy which achieved no evaluable disease. He maintained remission for ∼1 year after the first T_TCR-C4_ infusion. Subsequently, he relapsed extramedullary and was refractory to an additional T_TCR-C4_ infusion. Further details on this case and mechanisms of AML escape have been described previously.^2^

**Extended data Fig. 5.**
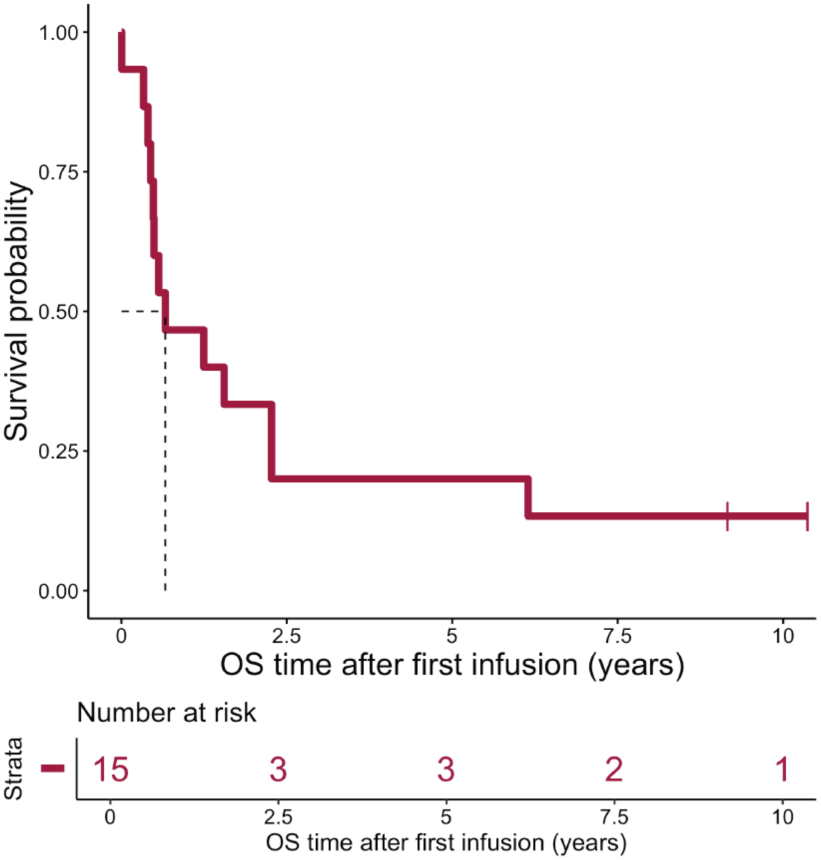
Overall survival of AML patients treated with T_TCRC4_ T cells. Kaplan Meier estimate of overall survival (OS) of 15 AML patients with evidence of disease post-HCT, who were subsequently treated with T_TCRC4_ T cells

**Extended data Fig. 6.**
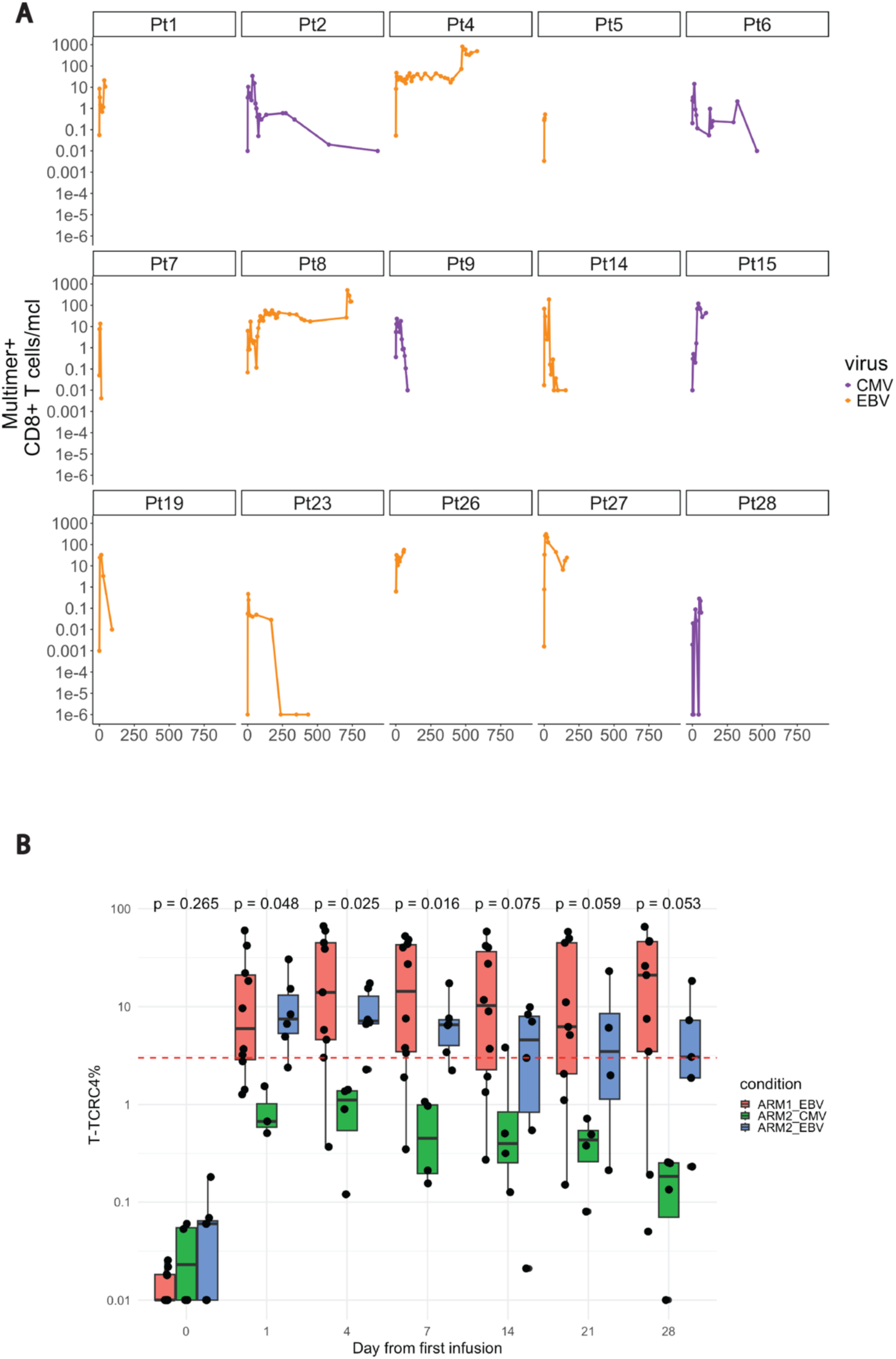
EBV specific vs. CMV specific T_TCR-C4_ persistence. **(A)** Line plots showing the absolute counts (cells/uL) in log scale of PB multimer+ CD8^+^ T cells collected after T_TCR-C4_ infusion and stratified by virus specificity. EBV-specific cells are represented in dark orange, and CMV-specific cells are represented in dark violet. Data points correspond to individual samples collected at different timepoints post-infusion. **(B)** Boxplots comparing the percentage of EBV-specific T_TCR-C4_ cells in Arm 1 (red) and Arm 2 (blue) with the percentage of CMV-specific T_TCR-C4_ cells in Arm 2 (green) across post-infusion timepoints. The y-axis indicates the percentage (log scale) of T_TCR-C4_ cells, while the x-axis represents the time post-infusion. Data were derived from flow-cytometry analysis. The horizontal dashed red line indicates the 3% threshold used to define persisting T_TCR-C4_ cells. The y-axis is displayed on a log10 scale. Statistical comparison between all three groups at each time point was performed using a Kruskal-Wallis test, with p-values displayed above the corresponding boxplots. Statistical significance was defined as p < 0.05.

**Extended data Fig. 7.**
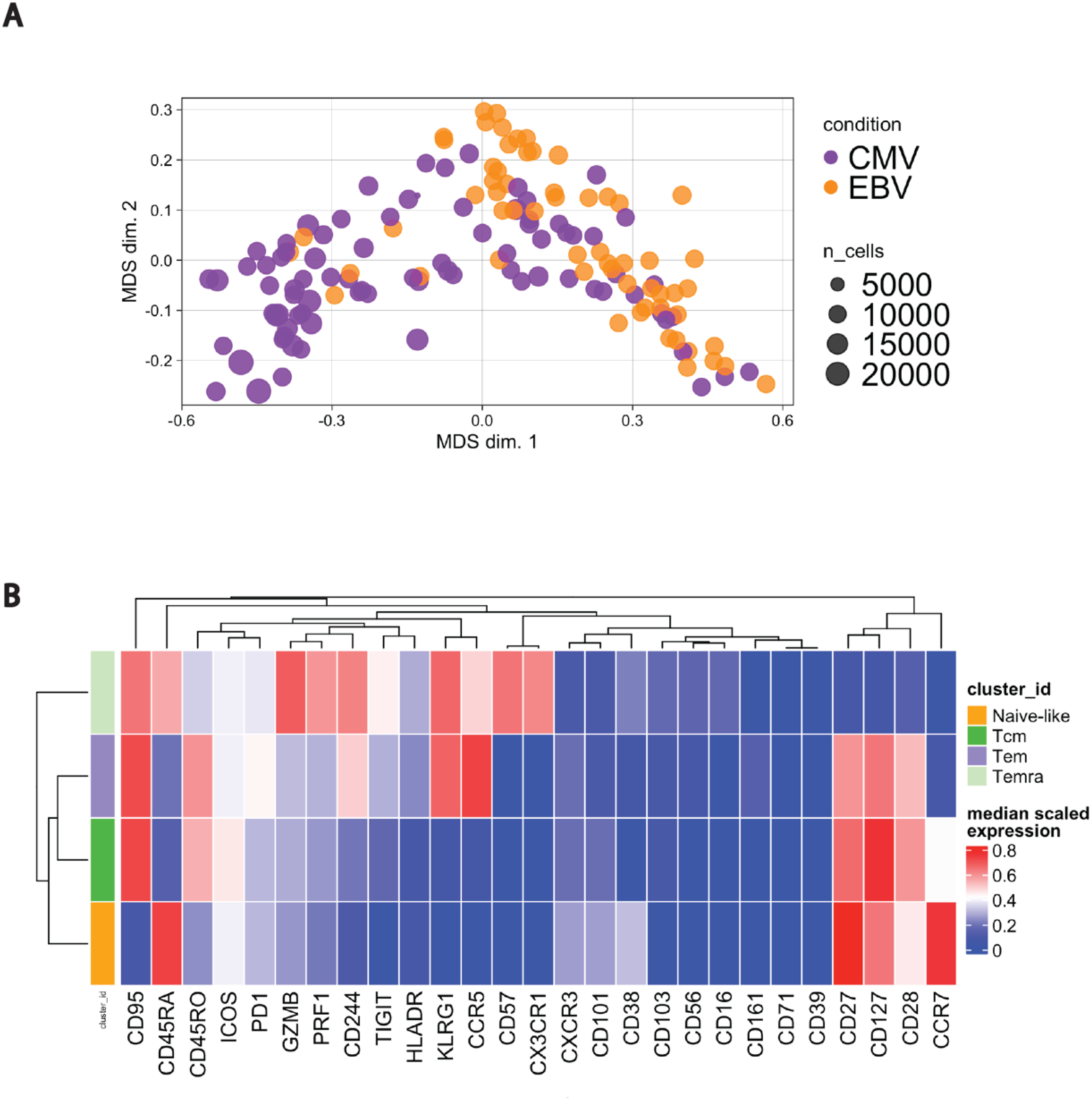
EBV and CMV specific CD8^+^ T-cell states. **(A)** Multidimensional (MDS) scaling plot illustrates the distribution of samples based on the expression of top varying markers, which are those markers showing the most variation in expression levels across the dataset. These markers are selected for their ability to highlight meaningful differences between samples or conditions. In this context, samples associated with CMV (violet dots) and EBV (orange dots) are plotted along the first two MDS dimensions (MDS dim. 1, MDS dim. 2). This unsupervised method visualizes sample clustering based on marker expression, with dissimilarities calculated using the median expression of markers across all cells in each sample. The plot provides an overview of variance and potential clustering patterns among the samples. **(B)** Heatmap illustrating the median expression levels of 27 markers across four PB CD8+ T cell subsets: Naïve-like (CD95-, CD45RA+, CCR7+, CD27+, CD28+), Effector Memory (Tem) (CD95+, CD45RA-, CD45RO+, CD27+, CD28+, CD127+), Central Memory (Tcm) (CD95+, CD45RA-, CD45RO+, CCR7+, CD27+, CD28+, CD127+), and Effector Memory RA+ (Temra) (CD95+, CD45RA+, CD45RO-, CD57+, KLRG1+). The data are scaled to normalize marker expression across the dataset before clustering, ensuring each marker contributes proportionally to cluster definitions while minimizing the impact of extreme values or outliers. Blue represents lower expression, and red indicates higher expression. Hierarchical clustering on the top dendrogram groups markers based on their expression patterns, revealing co-occurring markers across subsets, while the clustering on the left shows relationships between subsets based on their marker profiles, highlighting their similarities.

**Extended data Fig. 8.**
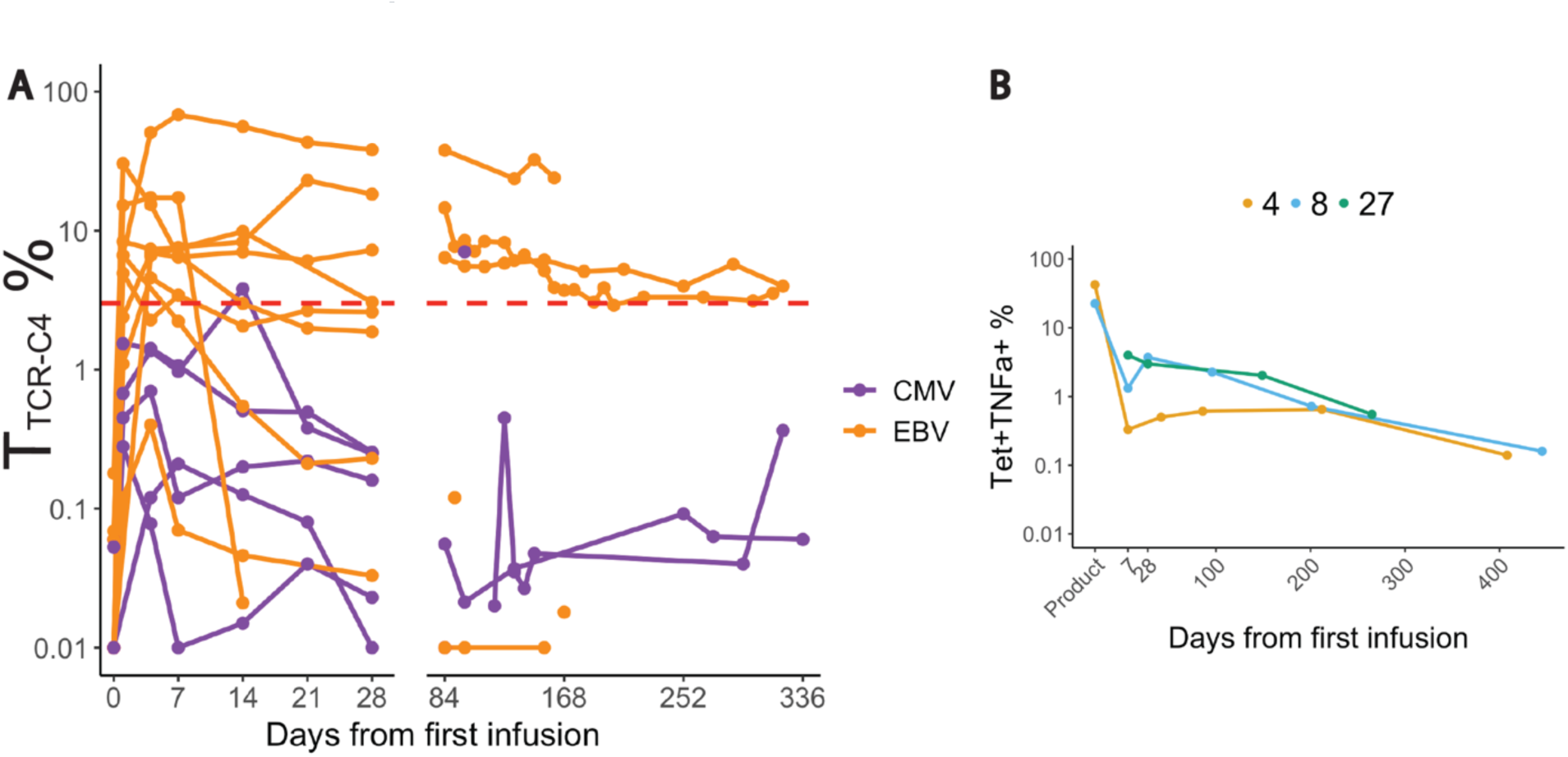
Persitence and functionality of T_TCR-C4_ post-first infusion. (A) Line plot showing the percentage (log scale) of T_TCR-C4_ in PBMCs collected after the first infusion across all patients (n =15), colored by virus-specificity (violet = CMV; orange = EBV), derived from flow- cytometry data **(B)** Line plot showing the TTCR_C4_^+^TNFα^+^ cells following WT1-peptide stimulation. The x axis represents the days from the first infusion, while the y axis indicates the percentage (log10 scale) of Tetramer^+^ TNFα^+^ gated on CD8^+^ T cells, based on flow cytometry data

**Extended data Fig. 9.**
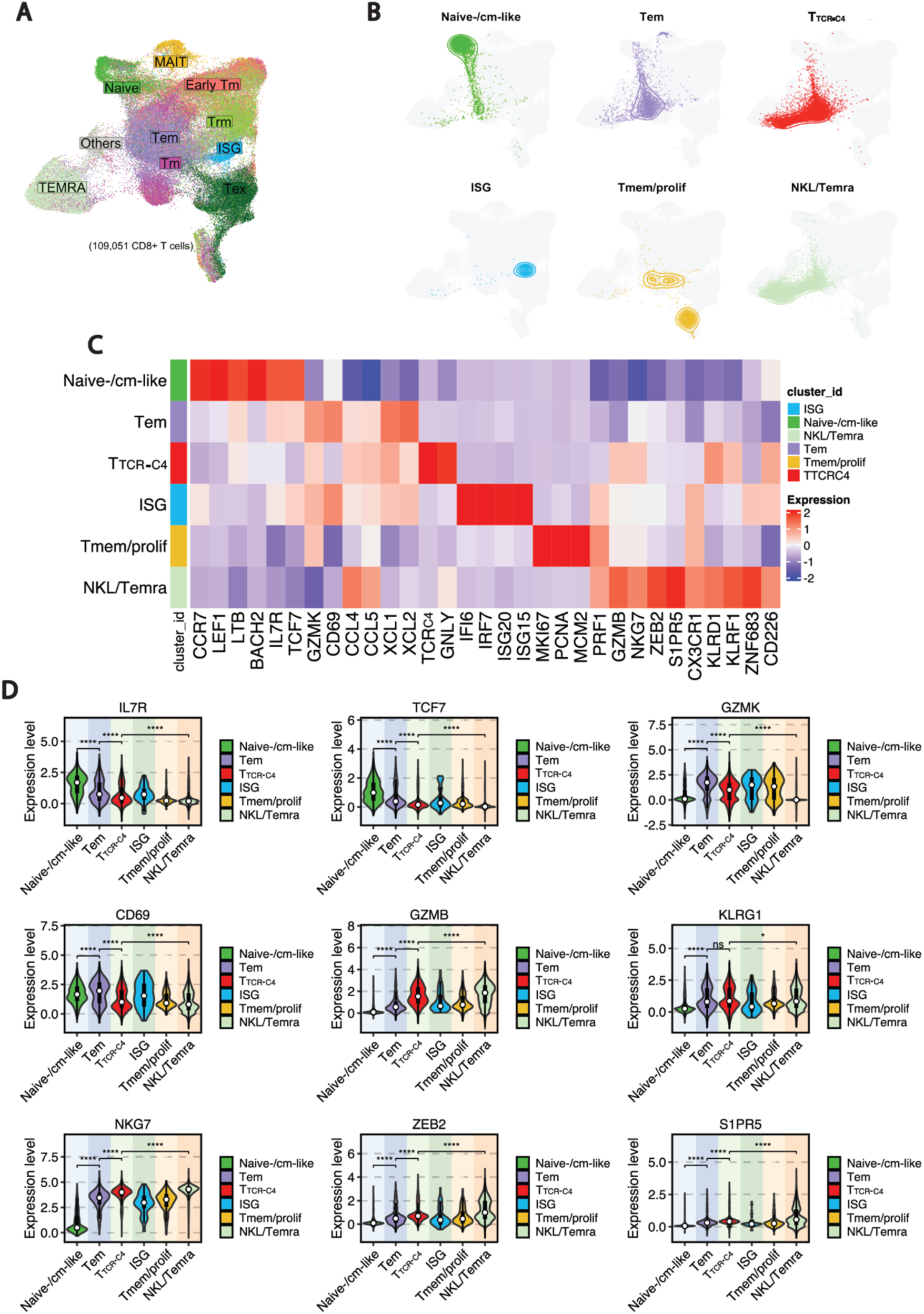
Identification of transcriptional endogenous and T_TCR-C4_ CD8^+^ T-cell states. **(A)** UMAP plot showing the distribution of CD8^+^ T-cell states from a published independent dataset^3^ used as a reference atlas. **(B)** The UMAP projections of each T-cell state defined in Fig. 3C are overlaid onto the reference atlas as contour plots. Contours corresponding to each subset from the query are color-coded as in Fig. 3C, allowing for easy visualization of the distribution of each subset within the reference atlas. **(C)** Heatmap showing the differential expression of manually curated genes (Supplemental Table 7) across the five annotated CD8^+^ T-cell subsets. Blue and red indicate the relative expression levels of each marker within each subset, with blue representing lower expression and red indicating higher expression. **(D)** Violin plots displaying the expression levels (y-axis) of stem-like (*IL7R*, *TCF7*), activation (*GZMK*, *CD69*), and cytotoxicity/NK-like (*GZMB*, *KLRG1*, *NKG7*, *ZEB2*, *S1PR5*) markers across CD8^+^ T-cell transcriptional states (x-axis). Statistical significance was assessed using the Wilcoxon rank sum test. Asterisks indicate the following thresholds of significance: *p < 0.05, **p < 0.01, ***p < 0.001, ****p < 0.0001.

**Extended data Fig. 10.**
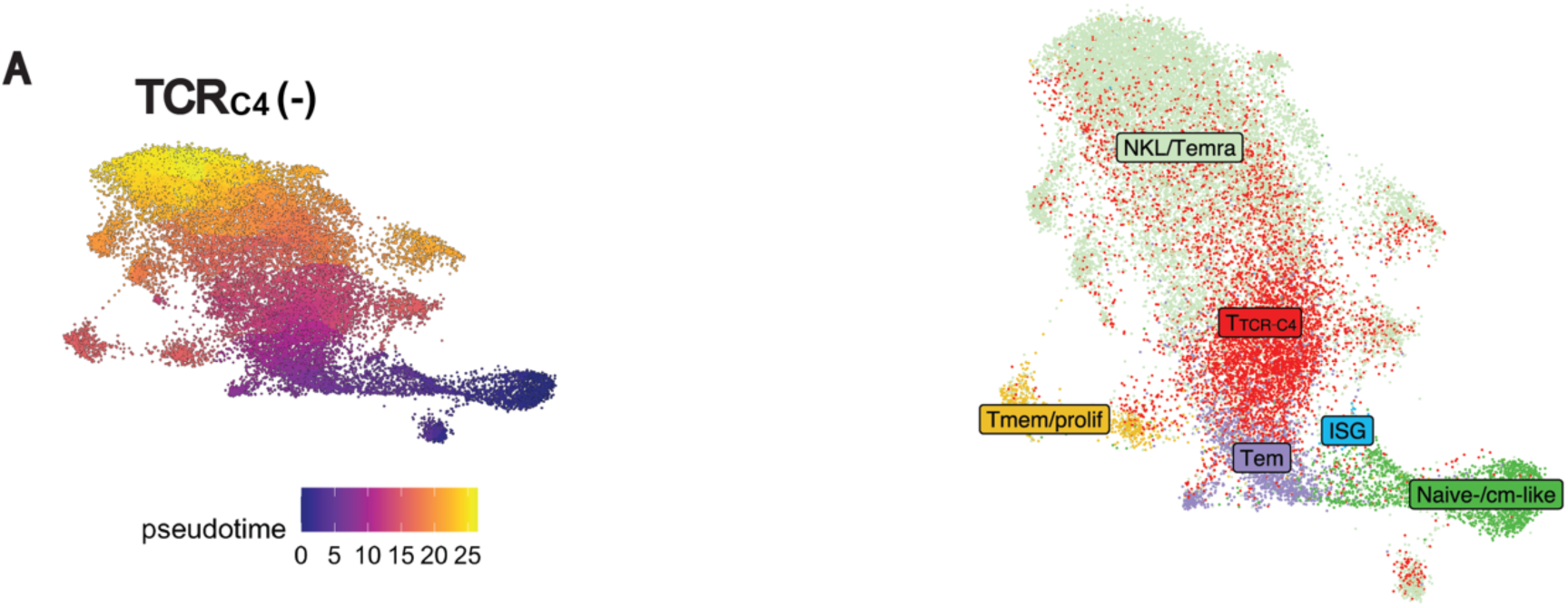
CD8^+^ transcriptional states after TCRC4 removal. **(A)** UMAP plot depicting the inferred developmental trajectory of CD8^+^ T-cell transcriptional states, as predicted by Monocle, following the removal of the TCRC4 transcript. The color scale, ranging from blu to orange/yellow, represents the pseudotime, where blue indicates earlier developmental stages and orange/yellow indicates later stages of the trajectory. **(B)** UMAP plot showing the distribution of CD8+ T-cell transcriptional states after TCRC4 removal, with the same coordinates as panel A. The T- cell subsets are labeled according to the clustering defined in Fig. 3D, providing context for how the subsets are positioned within the developmental trajectory in A.

**Extended data Fig. 11.**
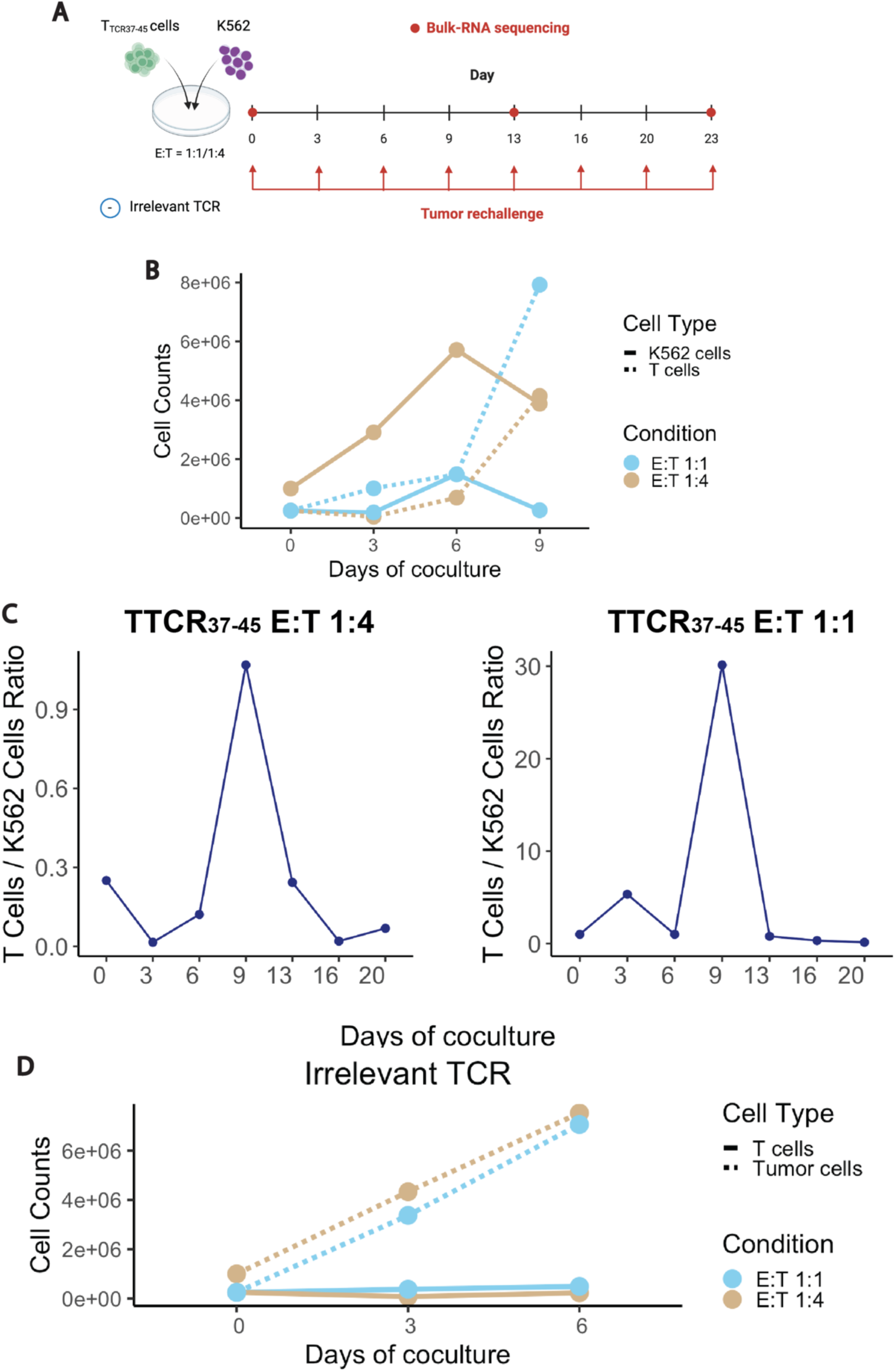
In-vitro dysfunction coculture system. **(A)** Experimental design of the in-vitro model used to study chronic tumor exposure of WT1-specific T cells (T_TCR37-45_ cells). The diagram shows two conditions with Effector (T_TCR37-45_) to Target (K562) (E:T) ratios of 1:1 and 1:4. Red arrows indicate tumor re-challenge every 3-4 days, while red circles on the timeline mark the time points when RNA extraction for bulk-RNA sequencing was performed. An irrelevant TCR-expressing T cell line was used as a negative control. The experiment involved exposing transgenic CD8^+^ WT1-specific T cells to HLA-A*0201-transduced K562 cells expressing high levels of WT1. This figure was realized using BioRender. **(B)** Line plots showing the changes in absolute cell counts (y-axis) over time (x-axis, days of coculture) for T_TCR37-45_ (solid lines) and K562 tumor cells (dashed lines) in coculture at E:T ratios of 1:1 and 1:4. The lines for each condition are colored sky blue (1:4 ratio) and tan (1:1 ratio). Cell counting was performed by flow cytometry. **(C)** Line plots showing the T cell to tumor cell ratio (y-axis) changes at the timepoints of tumor re-challenge (x-axis). Each plot represents one condition (left, E:T 1:4; right, E:T 1:1). Cell ratios were calculated based on flow cytometry data.**(D)** Line plots showing the changes in absolute cell counts (y-axis) over time (x-axis, days of coculture) for T cells with irrelevant TCR (solid lines) and K562 tumor cells (dashed lines) in coculture at E:T ratios of 1:1 and 1:4. The lines for each condition are colored sky blue (1:4 ratio) and tan (1:1 ratio). Cell counting was performed by flow cytometry.

**Extended data Fig. 12.**
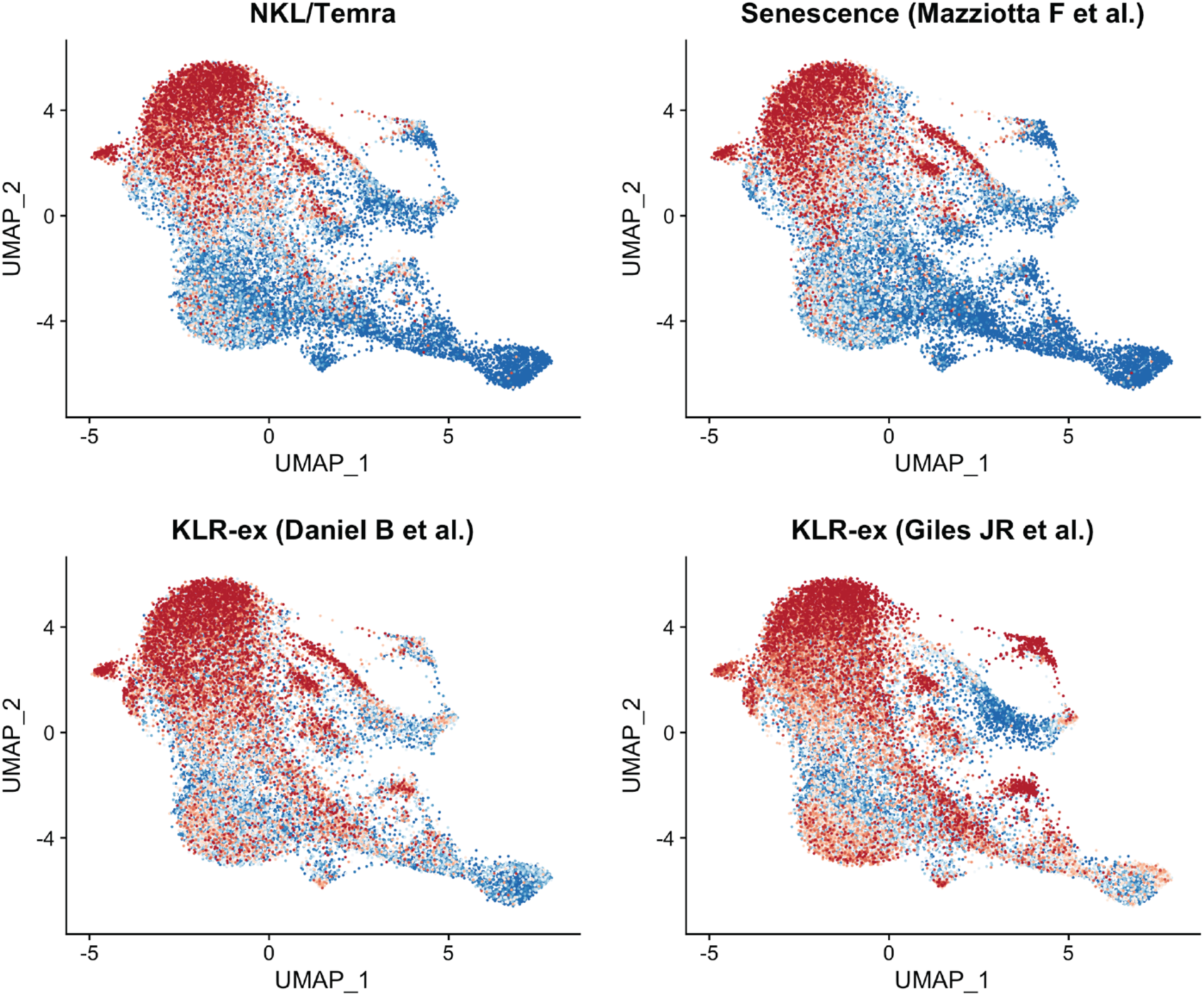
UMAP projection of KLR dysfunction signatures in CD8^+^ T cells. UMAP plots illustrate the spatial distribution of CD8^+^ T cells based on their expression of published KLR dysfunction gene signatures. ^4–6^ These signatures, derived from prior studies, were applied to our scRNAseq dataset to calculate a score for each cell, reflecting the degree of expression of the KLR dysfunction features. The scores are visualized using a gradient, where blue indicates low expression and red indicates high expression. Regions with high expression (red) align with the subset we identified as Temra NK-like cells, suggesting a functional and phenotypic overlap within this subset.

**Extended data Fig. 13.**
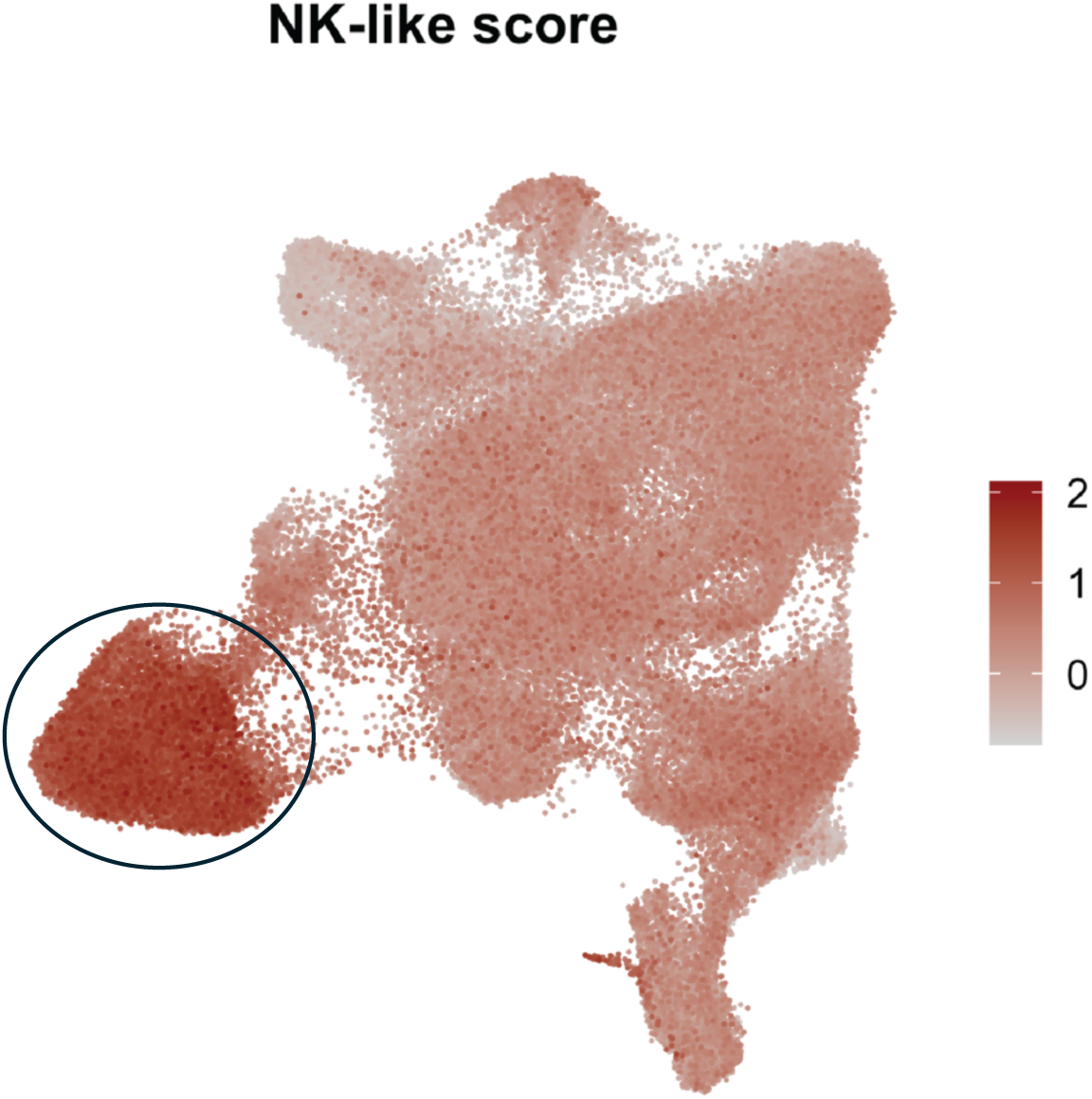
UMAP projection of NK-like module score based on gene expression in a tumor-infiltrating lymphocytes reference atlas. ^3^ UMAP plot projection showing the module score based on the expression of the following genes: *KLRG1*, *KLRD1*, *ZEB2*, *FCRL6*, *ADGRG1*, *S1PR5*, *FCGR3A*, *GZMB*, and *NKG7*. Each point represents an individual cell, and the color scale ranges from light grey (low module score) to dark red (high module score), reflecting the intensity of the gene set expression across cells. The black circle marks the area with the highest NK- like score, indicating the regions where the combined expression of NK-associated genes is most prominent.

**Extended data Fig. 14.**
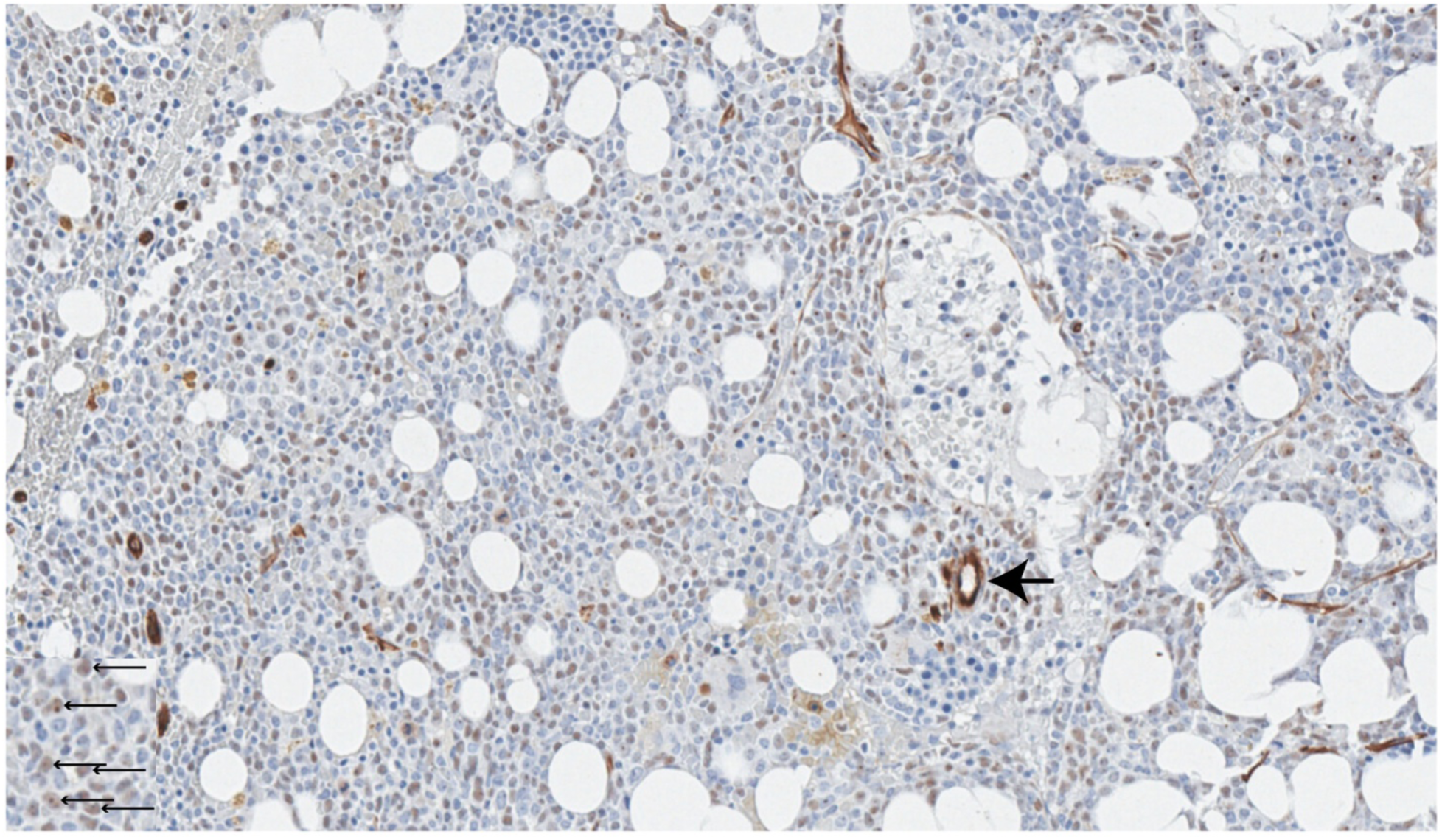
Immunohistochemistry of bone marrow biopsy stained for WT1. This immunohistochemistry image shows a bone marrow biopsy stained for WT1. Black arrows indicate WT1-positive nuclei, visible as distinct dark-stained areas within the cells, which signify expression of the WT1 protein.

**Extended data Fig. 15.**
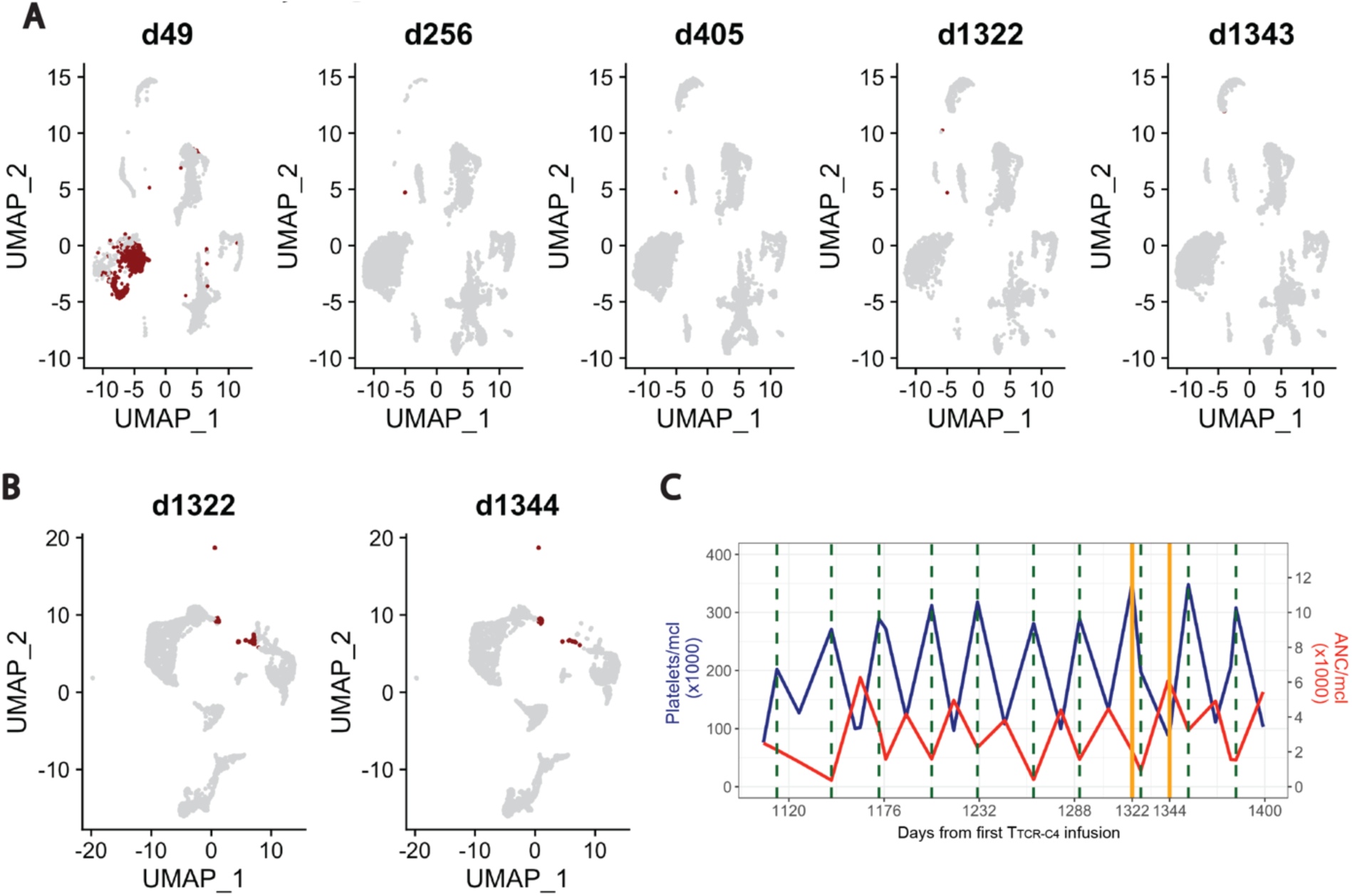
Patient 8 blasts and T cells characterization. **(A)** UMAP plot showing a blast score (dark red) calculated based on the co- expression of *CD34* and *XIST* (a female-specific gene) in a patient previously transplanted with a sex-mismatched donor (female patient, male donor). The score was used to identify cells expressing a high blast score. Each UMAP plot represents a different timepoint (day 49, 256, 405, 1322, 1343) after the first infusion, with lightgrey indicating low expression and dark red indicating high expression of the blast score. **(B)** UMAP plot showing a blast score calculated based on the co-expression of *XIST*, *CD34*, *CD33*, and *KIT*. This score was used to visualize cells with high blast scores in bone marrow samples. Each UMAP plot represents a specific timepoint (day 1322 and 1344) post-first T_TCR-C4_ infusion, **(C)** Line plot showing platelets and neutrophil absolute counts over time after the first infusion. Platelets (left y-axis, dark blue line plot, measured as platelets/μL × 1000) and neutrophils (right y-axis, red line plot, measured as ANC/μL × 1000) are shown in relation to the days after the first infusion (x-axis). Vertical dark green dashed lines indicate the timing of azacytidine cycles, while solid orange lines represent the timepoints when BM scRNA-seq was performed.

**Extended data Fig. 16.**
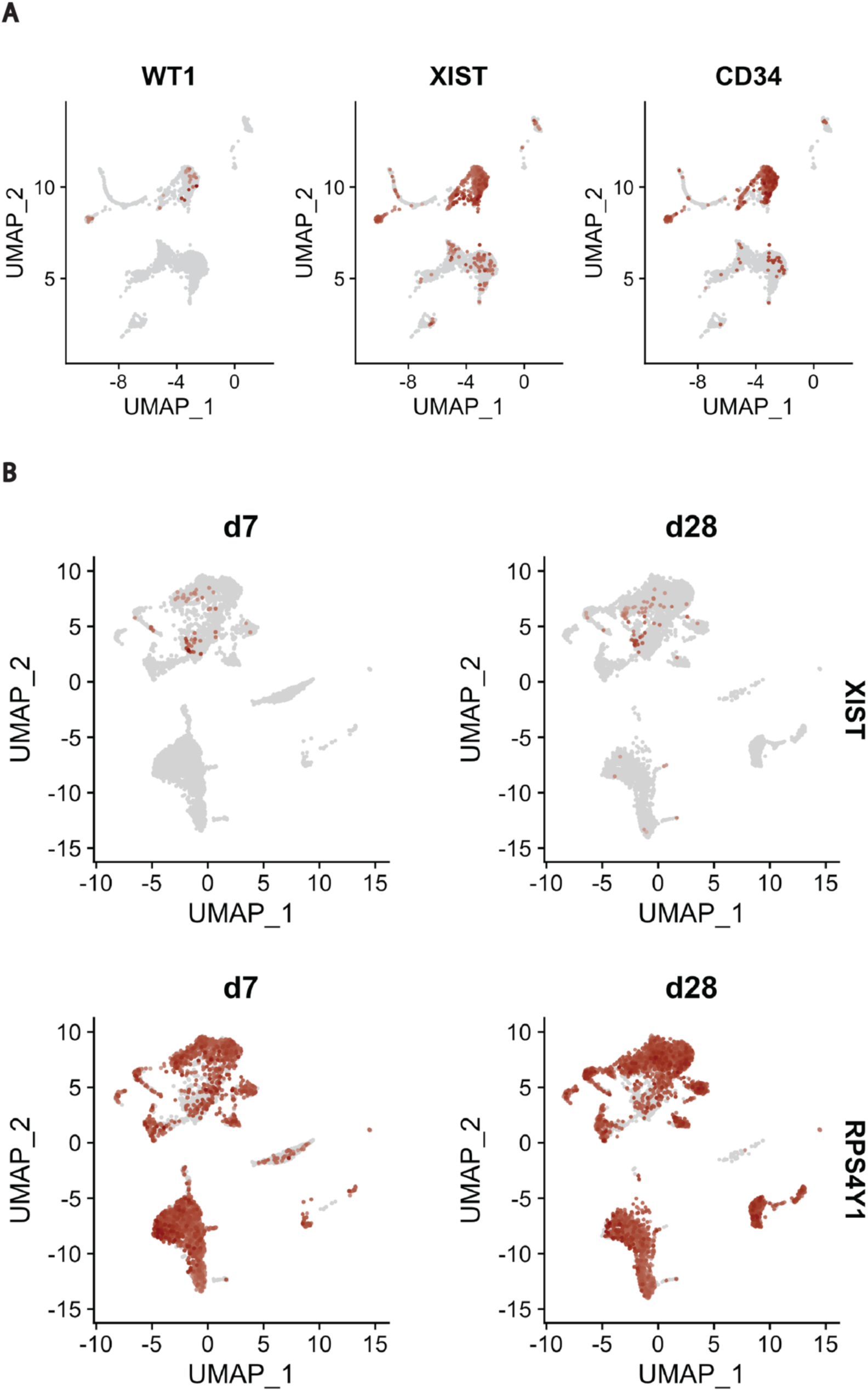
UMAP visualization of gene expression in AML patient samples. **(A)** UMAP plots at day 49 post-first T_TCR-C4_ infusion depict the expression patterns of leukemia-associated markers *WT1* and *XIST* (a female-specific gene) in patient 8, previously transplanted with a sex-mismatched donor (female patient, male donor). Additionally, *CD34* expression across all cell populations is shown. Cells with high expression of each marker are highlighted in red, while other cells appear in grey. **(B)** UMAP plots illustrate a longitudinal analysis of leukemia-associated gene expression in patient 26, who was transplanted with a sex-mismatched donor (female patient, male donor), at days 7 (d7) and 28 (d28) post-first T_TCR-C4_ infusion. *XIST* indicates leukemic cells, while *RPS4Y1* (a male- specific gene) marks non-leukemic cells. Cells with high gene expression are represented in dark red, and those with lower expression appear in light grey.

## References

1. Bejanyan, N., et al. Survival of patients with acute myeloid leukemia relapsing after allogeneic hematopoietic cell transplantation: a center for international blood and marrow transplant research study. Biol Blood Marrow Transplant 21, 454–459 (2015).

2. Schmid, C., et al. Treatment, risk factors, and outcome of adults with relapsed AML after reduced intensity conditioning for allogeneic stem cell transplantation. Blood 119, 1599–1606 (2012).

3. Webster, J.A., Luznik, L. & Gojo, I. Treatment of AML Relapse After Allo-HCT. Front Oncol 11, 812207 (2021).

4. Cheever, M.A., et al. The prioritization of cancer antigens: a national cancer institute pilot project for the acceleration of translational research. Clinical cancer research : an official journal of the American Association for Cancer Research 15, 5323–5337 (2009).

5. Inoue, K., et al. WT1 as a new prognostic factor and a new marker for the detection of minimal residual disease in acute leukemia. Blood 84, 3071–3079 (1994).

6. Menssen, H.D., et al. Presence of Wilms’ tumor gene (wt1) transcripts and the WT1 nuclear protein in the majority of human acute leukemias. Leukemia 9, 1060–1067 (1995).

7. Miyoshi, Y., et al. High expression of Wilms’ tumor suppressor gene predicts poor prognosis in breast cancer patients. Clinical cancer research : an official journal of the American Association for Cancer Research 8, 1167–1171 (2002).

8. Stromnes, I.M., Schmitt, T.M., Chapuis, A.G., Hingorani, S.R. & Greenberg, P.D. Re-adapting T cells for cancer therapy: from mouse models to clinical trials. Immunol Rev 257, 145–164 (2014).

9. Chapuis, A.G., et al. T cell receptor gene therapy targeting WT1 prevents acute myeloid leukemia relapse post-transplant. Nature Medicine 25, 1064–1072 (2019).

10. Kolb, H.J., et al. Graft-versus-leukemia effect of donor lymphocyte transfusions in marrow grafted patients. Blood 86, 2041–2050 (1995).

11. Kharfan-Dabaja, M.A., et al. Second allogeneic haematopoietic cell transplantation using HLA-matched unrelated versus T-cell replete haploidentical donor and survival in relapsed acute myeloid leukaemia. Br J Haematol 193, 592–601 (2021).

12. Fraietta, J.A., et al. Determinants of response and resistance to CD19 chimeric antigen receptor (CAR) T cell therapy of chronic lymphocytic leukemia. Nat Med 24, 563–571 (2018).

13. Deng, Q., et al. Characteristics of anti-CD19 CAR T cell infusion products associated with efficacy and toxicity in patients with large B cell lymphomas. Nat Med 26, 1878–1887 (2020).

14. Kirouac, D.C., et al. Author Correction: Deconvolution of clinical variance in CAR-T cell pharmacology and response. Nat Biotechnol 41, 1655 (2023).

15. Fraietta, J.A., et al. Disruption of TET2 promotes the therapeutic efficacy of CD19-targeted T cells. Nature 558, 307–312 (2018).

16. Eyquem, J., et al. Targeting a CAR to the TRAC locus with CRISPR/Cas9 enhances tumour rejection. Nature 543, 113–117 (2017).

17. Blank, C.U., et al. Defining ’T cell exhaustion’. Nat Rev Immunol 19, 665–674 (2019).

18. Grosser, R., Cherkassky, L., Chintala, N. & Adusumilli, P.S. Combination Immunotherapy with CAR T Cells and Checkpoint Blockade for the Treatment of Solid Tumors. Cancer Cell 36, 471–482 (2019).

19. Hirayama, A.V., et al. Timing of anti-PD-L1 antibody initiation affects efficacy/toxicity of CD19 CAR T-cell therapy for large B-cell lymphoma. Blood Adv 8, 453–467 (2024).

20. Penter, L., et al. Mechanisms of response and resistance to combined decitabine and ipilimumab for advanced myeloid disease. Blood (2023).

21. Rutella, S., et al. Immune dysfunction signatures predict outcomes and define checkpoint blockade-unresponsive microenvironments in acute myeloid leukemia. The Journal of clinical investigation 132(2022).

22. Mazziotta, F., et al. CD8+ T-cell Differentiation and Dysfunction Inform Treatment Response in Acute Myeloid Leukemia. Blood (2024).

23. Abbas, H.A., et al. Single cell T cell landscape and T cell receptor repertoire profiling of AML in context of PD-1 blockade therapy. Nature communications 12, 6071–6071 (2021).

24. Zeidner, J.F., et al. Phase II Trial of Pembrolizumab after High-Dose Cytarabine in Relapsed/Refractory Acute Myeloid Leukemia. Blood cancer discovery 2, 616–629 (2021).

25. Penter, L. & Wu, C.J. Therapy response in AML: a tale of two T cells. Blood 144, 1134–1136 (2024).

26. Döhner, H., et al. Diagnosis and management of AML in adults: 2017 ELN recommendations from an international expert panel. Blood 129, 424–447 (2017).

27. Schmid, C., et al. Donor lymphocyte infusion in the treatment of first hematological relapse after allogeneic stem-cell transplantation in adults with acute myeloid leukemia: a retrospective risk factors analysis and comparison with other strategies by the EBMT Acute Leukemia Working Party. J Clin Oncol 25, 4938–4945 (2007).

28. Berger, C., et al. Adoptive transfer of effector CD8+ T cells derived from central memory cells establishes persistent T cell memory in primates. J Clin Invest 118, 294–305 (2008).

29. Schmidt, F., et al. In-depth analysis of human virus-specific CD8(+) T cells delineates unique phenotypic signatures for T cell specificity prediction. Cell Rep 42, 113250 (2023).

30. Quintelier, K., et al. Analyzing high-dimensional cytometry data using FlowSOM. Nat Protoc 16, 3775–3801 (2021).

31. Larbi, A. & Fulop, T. From “truly naïve” to “exhausted senescent” T cells: When markers predict functionality. Cytometry Part A 85, 25–35 (2014).

32. Gattinoni, L., Speiser, D.E., Lichterfeld, M. & Bonini, C. T memory stem cells in health and disease. Nature Medicine 23, 18–27 (2017).

33. Wherry, E.J. & Kurachi, M. Molecular and cellular insights into T cell exhaustion. Nat Rev Immunol 15, 486–499 (2015).

34. Andreatta, M., Berenstein, A.J. & Carmona, S.J. scGate: marker-based purification of cell types from heterogeneous single-cell RNA-seq datasets. Bioinformatics (Oxford, England) 38, 2642–2644 (2022).

35. Zheng, L., et al. Pan-cancer single-cell landscape of tumor-infiltrating T cells. Science (New York, N.Y.) 374, abe6474-abe6474 (2021).

36. Szabo, P.A., et al. Single-cell transcriptomics of human T cells reveals tissue and activation signatures in health and disease. Nature communications 10, 4706–4706 (2019).

37. Koh, J.-Y., et al. Identification of a distinct NK-like hepatic T-cell population activated by NKG2C in a TCR-independent manner. Journal of Hepatology 77, 1059–1070 (2022).

38. Daniel, B., et al. Divergent clonal differentiation trajectories of T cell exhaustion. Nature immunology 23, 1614–1627 (2022).

39. Giles, J.R., et al. Shared and distinct biological circuits in effector, memory and exhausted CD8+ T cells revealed by temporal single-cell transcriptomics and epigenetics. Nature immunology 23, 1600–1613 (2022).

40. Trapnell, C., et al. The dynamics and regulators of cell fate decisions are revealed by pseudotemporal ordering of single cells. Nature Biotechnology 32, 381–386 (2014).

41. Bergen, V., Lange, M., Peidli, S., Wolf, F.A. & Theis, F.J. Generalizing RNA velocity to transient cell states through dynamical modeling. Nature biotechnology 38, 1408–1414 (2020).

42. Good, C.R., et al. An NK-like CAR T cell transition in CAR T cell dysfunction. Cell 184, 6081–6100.e6026 (2021).

43. Lahman, M.C., et al. Targeting an alternate Wilms’ tumor antigen 1 peptide bypasses immunoproteasome dependency. Science translational medicine 14, eabg8070-eabg8070 (2022).

44. Dufva, O., et al. Immunogenomic Landscape of Hematological Malignancies. Cancer cell 38, 424–428 (2020).

45. Lasry, A., et al. An inflammatory state remodels the immune microenvironment and improves risk stratification in acute myeloid leukemia. Nature Cancer (2022).

46. Schalck, A., et al. Single-Cell Sequencing Reveals Trajectory of Tumor-Infiltrating Lymphocyte States in Pancreatic Cancer. Cancer Discov 12, 2330–2349 (2022).

47. Zhang, C., et al. A single-cell analysis reveals tumor heterogeneity and immune environment of acral melanoma. Nat Commun 13, 7250 (2022).

48. Wu, F., et al. Single-cell profiling of tumor heterogeneity and the microenvironment in advanced non-small cell lung cancer. Nat Commun 12, 2540 (2021).

49. Craddock, C., et al. Clinical activity of azacitidine in patients who relapse after allogeneic stem cell transplantation for acute myeloid leukemia. Haematologica 101, 879–883 (2016).

50. Jackson, S.E., Sedikides, G.X., Okecha, G. & Wills, M.R. Generation, maintenance and tissue distribution of T cell responses to human cytomegalovirus in lytic and latent infection. Med Microbiol Immunol 208, 375–389 (2019).

51. Pociupany, M., Snoeck, R., Dierickx, D. & Andrei, G. Treatment of Epstein-Barr Virus infection in immunocompromised patients. Biochem Pharmacol 225, 116270 (2024).

52. Appay, V., et al. Memory CD8+ T cells vary in differentiation phenotype in different persistent virus infections. Nat Med 8, 379–385 (2002).

53. Newell, E.W., et al. Combinatorial tetramer staining and mass cytometry analysis facilitate T-cell epitope mapping and characterization. Nat Biotechnol 31, 623–629 (2013).

54. Catalina, M.D., Sullivan, J.L., Brody, R.M. & Luzuriaga, K. Phenotypic and functional heterogeneity of EBV epitope-specific CD8+ T cells. J Immunol 168, 4184–4191 (2002).

55. Callan, M.F., et al. CD8(+) T-cell selection, function, and death in the primary immune response in vivo. J Clin Invest 106, 1251–1261 (2000).

56. Abbott, R.J., et al. Asymptomatic Primary Infection with Epstein-Barr Virus: Observations on Young Adult Cases. J Virol 91(2017).

57. Young, L.S. & Rickinson, A.B. Epstein-Barr virus: 40 years on. Nat Rev Cancer 4, 757–768 (2004).

58. Sinzger, C., et al. Fibroblasts, epithelial cells, endothelial cells and smooth muscle cells are major targets of human cytomegalovirus infection in lung and gastrointestinal tissues. J Gen Virol 76 (Pt **4**), 741–750 (1995).

59. Nakamura, K. & Smyth, M.J. Myeloid immunosuppression and immune checkpoints in the tumor microenvironment. Cell Mol Immunol 17, 1–12 (2020).

60. Muroyama, Y. & Wherry, E.J. Memory T-Cell Heterogeneity and Terminology. Cold Spring Harbor Perspectives in Biology 13, a037929–a037929 (2021).

61. Desai, P.N., et al. Single-Cell Profiling of CD8+ T Cells in Acute Myeloid Leukemia Reveals a Continuous Spectrum of Differentiation and Clonal Hyperexpansion. *Cancer Immunol Res*, OF1-OF18 (2023).

62. Schuh, A.C., et al. Azacitidine in adult patients with acute myeloid leukemia. Crit Rev Oncol Hematol 116, 159–177 (2017).

63. Goodyear, O., et al. Induction of a CD8+ T-cell response to the MAGE cancer testis antigen by combined treatment with azacitidine and sodium valproate in patients with acute myeloid leukemia and myelodysplasia. Blood 116, 1908–1918 (2010).

64. Costantini, B., et al. The effects of 5-azacytidine on the function and number of regulatory T cells and T-effectors in myelodysplastic syndrome. Haematologica 98, 1196–1205 (2013).

65. El Khawanky, N., et al. Demethylating therapy increases anti-CD123 CAR T cell cytotoxicity against acute myeloid leukemia. Nat Commun 12, 6436 (2021).

66. Hahne, F., et al. flowCore: a Bioconductor package for high throughput flow cytometry. BMC bioinformatics 10, 106–106 (2009).

67. Nowicka, M., et al. CyTOF workflow: differential discovery in high-throughput high- dimensional cytometry datasets. F1000Research 6, 748–748 (2017).

68. Lun, A.T.L., et al. EmptyDrops: distinguishing cells from empty droplets in droplet-based single-cell RNA sequencing data. Genome biology 20, 63–63 (2019).

69. Dobin, A., et al. STAR: ultrafast universal RNA-seq aligner. Bioinformatics 29, 15–21 (2013).

70. Amezquita, R.A., et al. Orchestrating single-cell analysis with Bioconductor. Nature methods 17, 137–145 (2020).

71. Bais, A.S. & Kostka, D. scds: computational annotation of doublets in single-cell RNA sequencing data. Bioinformatics (Oxford, England) 36, 1150–1158 (2020).

72. McCarthy, D.J., Campbell, K.R., Lun, A.T.L. & Wills, Q.F. Scater: pre-processing, quality control, normalization and visualization of single-cell RNA-seq data in R. *Bioinformatics (Oxford*, England) 33, 1179–1186 (2017).

73. Stuart, T., et al. Comprehensive Integration of Single-Cell Data. Cell 177, 1888–1902.e1821 (2019).

74. Butler, A., Hoffman, P., Smibert, P., Papalexi, E. & Satija, R. Integrating single-cell transcriptomic data across different conditions, technologies, and species. Nature biotechnology 36, 411–420 (2018).

75. Gu, Z., Eils, R. & Schlesner, M. Complex heatmaps reveal patterns and correlations in multidimensional genomic data. Bioinformatics 32, 2847–2849 (2016).

76. Andreatta, M., et al. Interpretation of T cell states from single-cell transcriptomics data using reference atlases. Nature Communications 12, 2965–2965 (2021).

77. Wolf, F.A., Angerer, P. & Theis, F.J. SCANPY: large-scale single-cell gene expression data analysis. Genome biology 19, 15–15 (2018).

78. Borcherding, N., Bormann, N.L. & Kraus, G. scRepertoire: An R-based toolkit for single-cell immune receptor analysis. F1000Research 9, 47–47 (2020).

79. Trapnell, C., et al. The dynamics and regulators of cell fate decisions are revealed by pseudotemporal ordering of single cells. Nat Biotechnol 32, 381–386 (2014).

80. Love, M.I., Huber, W. & Anders, S. Moderated estimation of fold change and dispersion for RNA-seq data with DESeq2. Genome Biology 15, 550–550 (2014).

## References

1. Chapuis, A.G., et al. T cell receptor gene therapy targeting WT1 prevents acute myeloid leukemia relapse post-transplant. Nature Medicine 25, 1064–1072 (2019).

2. Lahman, M.C., et al. Targeting an alternate Wilms’ tumor antigen 1 peptide bypasses immunoproteasome dependency. Science translational medicine 14, eabg8070- eabg8070 (2022).

3. Zheng, L., et al. Pan-cancer single-cell landscape of tumor-infiltrating T cells. Science (New York, N.Y.) 374, abe6474-abe6474 (2021).

4. Mazziotta, F., et al. CD8+ T-cell Differentiation and Dysfunction Inform Treatment Response in Acute Myeloid Leukemia. Blood (2024).

5. Daniel, B., et al. Divergent clonal differentiation trajectories of T cell exhaustion. Nature immunology 23, 1614–1627 (2022).

6. Giles, J.R., et al. Shared and distinct biological circuits in effector, memory and exhausted CD8+ T cells revealed by temporal single-cell transcriptomics and epigenetics. Nature immunology 23, 1600–1613 (2022).

