## Supplementary tables for "Acute Myeloid Leukemia Skews Therapeutic WT1-specific CD8 TCR-T Cells Towards an NK-like Phenotype that Compromises Function and Persistence"

#### S. Table 1

| Patients characteritics | | |
|---|---|---|
| Age, years | Median | 40 |
| | Range | 3 - 74 |
| Gender | Male | 40% |
| | Female | 60% |
| ELN risk stratification | Favorable/Intermediate | 53% |
| | Adverse | 47% |
| Other AML characteristics | Secondary or treatment-related | 27% |
| $T_{TCRC4}$ virus-specificity | CMV | 33% |
| | EBV | 67% |
| Number of allo-HCT before $T_{TCRC4}$ infusion | 1 | 60% |
| | 2 | 40% |
| Disease status at 1st $T_{TCRC4}$ infusion | $CR_{MRD-}$ | 53% |
| | $CR_{MRD+}$ | 33% |
| | Refractory | 13% |
| Lymphodepletion | y | 20% |
| | n | 80% |
| GVHD after $T_{TCRC4}$ infusion | aGVHD | 13% |
| | cGVHD | 20% |

### S. Table 2

| NCI CTCAE v4.0* | | Definitely/probably associated with TTCR-C4 | | Unrelated/unlikely** associated with TTCR-C4 | | |
|---|---|---|---|---|---|---|
| | | Grade 3 | Grade 4 | Grade 3 | Grade 4 | Grade 5 |
| Cytokine-mediated | Cytokine Release Syndrome (fevers, chills, rigors, nausea) | 2 | | 1 | | |
| Blood and lymphatic system disorders | Lymphopenia | 5 | 2 | 3 | 3 | |
| | Thrombocytopenia | 5 | 2 | 7 | 6 | |
| | Neutrophil count decreased | 8 | 1 | 12 | 9 | |
| | Anemia | 2 | | 15 | | |
| GVHD-associated | Anorexia | | | 1 | | |
| | Diarrhea | | | 1 | | |
| | Maculo-papular rash | 1 | | 3 | | |
| | Hypoxia | 1 | | 1 | | |
| | Reduced joint range of motion | | | 1 | | |
| | Skin sclerosis | | | 1 | | |
| Metabolic/renal disorders | Hypokalemia | | | 2 | 1 | |
| | Hyponatremia | | | 10 | | |
| | Hypophosphatemia | | | 4 | | |
| | Proteinuria | | | 1 | | |
| | Hypocalcemia | | | 2 | | |
| | Hypoalbuminemia | | | 1 | | |
| | Hyperglycemia | | | 7 | | |
| | Chronic Kidney Disease | | | 1 | | |
| Miscellaneous | Infection | | | 6 | 1 | |
| | Hypothension | 2 | | 1 | | |
| | Hypertension | 2 | | | | |
| | Peripheral motor neuropathy | | | 1 | | |
| | Fatigue | | | 2 | | |
| | Duodenal Obstruction | | | 1 | | |
| | Abdominal pain | | | 1 | | |
| | Rectal Pain | 1 | | | | |
| | Nausea | | | 1 | | |
| | Aspartate Aminotransferase increased | 1 | | | 1 | |
| | Mucositis oral | | | 2 | | |
| | Dry Mouth | | | 1 | | |
| | Pulmonary Edema | | | | 1 | |
| | Multi-organ Failure | | | | | 1 |
| | Alopecia | | | | 1 | |
| | Thromboembolic event | | | | 1 | |
| | Activated partial thromboplastin time prolonged | | | 1 | | |
| | Sinus tachycardia | | | 1 | | |

***National Cancer Institute Common Terminology Criteria for Adverse Events version 4.0; **Adverse events recorded as having definite or probable association with TTCR-C4 were considered related**

### S. Table 3

| Spectral Flow-cytometry panel | | |
|---|---|---|
| **Marker** | Clone | Fluorescent Tag |
| **TIGIT** | A15153G | BV421 |
| **CD197 (CCR7)** | 3D12 | BB700 |
| **Ki-67** | B56 | BV480 |
| **CD57** | NK-1 | BUV395 |
| **CD45RA** | HI100 | BV570 |
| **CD27** | O323 | BV605 |
| **CD95 (FAS)** | DX29 | BV650 |
| **CD127 (IL-7Ra)** | A019D5 | BV711 |
| **CD28** | CD28.2 | BV785 |
| **T-Bet** | 4B10 | KIRAVIA Blue 520 |
| **CD8** | RPA-T8 | BUV737 |
| **TCF1 (TCF7)** | C63D9 | PE |
| **CD279 (PD-1)** | EH12.2H7 | PE-Dazzle 594 |
| **CD69** | FN50 | PE-Cy5 |
| **Granulysin** | DH2 | PE-Cy7 |
| **KLRG1** | Z7-205.rMAb | PerCP-eFluor 710 |
| **Granzyme K** | G3H69 | RB780 |
| **Tetramer** | | APC |
| **Granzyme B** | GB11 | Alexa Fluor 700 |
| **CD38** | HIT2 | APC-Fire 810 |
| **CD3** | UCHT1 | APC-Fire 750 |
| **CD366 (TIM3)** | F38-2E2 | PE-Fire 810 |
| **Viability** | | LD Blue |
| **CD33 (DUMP)** | WM53 | BV510 |
| **CD14 (DUMP)** | 63D3 | BV510 |

### S. Table 4

| | Days after TTCRC4 infusion | | | |
|---|---|---|---|---|
| IDs | T1 | T2 | T3 | T4 |
| 126 | 1 | 7 | 28 | 117 |
| 90 | 1 | 7 | 28 | 110 |
| 683 | 3 | 8 | 28 | 149 |

### S. Table 5

| IDs | Days after first infusion | | | | | Arm |
|---|---|---|---|---|---|---|
| 10 | D320 | | | | | Prophylactic |
| 18 | D253 | | | | | Prophylactic |
| 4 | D100 | D581 | | | | Treatment |
| 8 | D49 | D265 | D405 | D1322 | D1343 | Treatment |
| 15 | D97 | | | | | Treatment |
| 26 | D7 | D28 | | | | Treatment |
| 27 | D21 | D58 | D178 | | | Treatment |

### S. Table 6

| naïve-like | activation, memory | TTCR-C4 | interferon signaling genes | proliferation, memory | NK-like |
|---|---|---|---|---|---|
| CCR7 | CD69 | TCRC4 | ISG15 | MKI67 | KLRF1 |
| SELL | TIGIT | | ISG20 | MCM5 | KIR3DL1 |
| TCF7 | GZMK | | IRF7 | MCM7 | NKG7 |
| LEF1 | IL7R | | IFI6 | CD27 | FCGR3A |
| IL7R | CXCR4 | | | | GZMB |
| | | | | | KLRD1 |
| | | | | | PRF1 |

### S. Table 7

| Naïve-like | Tem | ISG | Tmem-prolif | NKL/TEMRA |
|---|---|---|---|---|
| CCR7 | GZMK | IFI6 | MKI67 | GNLY |
| LEF1 | CD69 | IRF7 | PCNA | PRF1 |
| LTB | CCL4 | ISG20 | MCM2 | GZMB |
| BACH2 | CCL5 | ISG15 | | NKG7 |
| IL7R | XCL1 | | | ZEB2 |
| TCF7 | XCL2 | | | S1PR5 |
| | | | | CXrCR1 |
| | | | | KLRD1 |
| | | | | KLRF1 |
| | | | | ZNF683 |
| | | | | CD226 |

### S. Table 8

| ID | Days after 1st infusion | Status | Blasts |
|---|---|---|---|
| 10 | 320 | AML- | No blasts |
| 18 | 253 | AML- | No blasts |
| 4 | 100 | AML- | No blasts |
| | 581 | AML+ | |
| 8 | 49 | AML+ | |
| | 265 | AML- | No blasts |
| | 405 | AML+ | 12.50% |
| | 1322 | AML+ | 0.30% |
| | 1343 | AML+ | 0.20% |
| 15 | 97 | AML+ | 9.80% |
| 26 | D7 | AML+ | |
| | D28 | AML+ | 9% |
| 27 | 21 | AML- | No blasts |
| | 58 | AML- | No blasts |
| | 178 | AML+ | 11% |

| | |
|---|---|
| (red) | Blasts identified via scRNAseq (Supplemental Figure 16) |
| (green) | Described in Lahman, M.C., et al. Targeting an alternate Wilms' tumor antigen 1 peptide bypasses immunoproteasome dependency. Science translational medicine 14, eabg8070-eabg8070 (2022). |

### S. Table 9

| Naïve-Like | Tem | TTCRC4 | ISG | Tmem/prolif | NKL/Temra | exhaustion |
|---|---|---|---|---|---|---|
| LTB | GZMK | TCRC4 | MX1 | HMOX1 | GZMH | PDCD1 |
| SELL | DUSP2 | GNLY | ISG15 | GAPDH | ZEB2 | LAG3 |
| NOSIP | ZNF331 | PTMS | IFI6 | STMN1 | NKG7 | HAVCR2 |
| LEF1 | NR4A2 | YBX3 | STAT1 | ACTG1 | TTC38 | CTLA4 |
| LEPROTL1 | FOS | TRBV14 | IRF7 | CARHSP1 | S1PR5 | TIM3 |
| IL7R | CSRNP1 | CMC1 | EIF2AK2 | MCM5 | GZMB | LAG3 |
| TCF7 | ZFP36L2 | HOPX | PARP9 | H2AFV | CST7 | CD160 |
| CCR7 | JUNB | TIGIT | IFITM1 | CACYBP | RAP1B | BTLA |
| PIK3IP1 | CXCR4 | RNF19A | MX2 | ANP32B | SYNGR1 | ENTPD1 |
| LDHB | RGS1 | LAG3 | BST2 | HMGB2 | HLA-DRB5 | |
| TMEM123 | DNAJB1 | UCP2 | LGALS9 | COX5A | DIP2A | |
| SERINC5 | PTGER4 | CCL5 | TRIM22 | S1PR4 | S100A4 | |
| NELL2 | IFNGR1 | FGFBP2 | CD28 | NUDT21 | FLNA | |
| PIM1 | CD27 | FDFT1 | UBE2L6 | HLA-DQB1 | KLRD1 | |
| FOXP1 | DUSP1 | PDE4D | EPSTI1 | HMGN2 | HLA-DRB1 | |
| CAMK4 | PIK3R1 | ALOX5AP | LY6E | HLA-DMA | EMP3 | |
| PABPC1 | RHOH | CD6 | IFI35 | RAD21 | HLA-DPB1 | |
| PASK | BTG1 | S100A10 | CHMP5 | CNN2 | YWHAQ | |
| BCL2 | SLC2A3 | H2AFZ | OAS1 | TFDP1 | CLIC3 | |
| RCAN3 | TSPYL2 | KLF3 | SAMD9L | TUBA1B | ANXA1 | |
| OXNAD1 | CREM | CLDND1 | SAT1 | HLA-DRA | C12orf75 | |
| SARAF | ZFP36 | PPP1R18 | XAF1 | SAE1 | GSTP1 | |
| FLT3LG | PPP1R15A | MARCKSL1 | TAP1 | SIRPG | BATF | |
| DGKA | PDE4B | ADGRG1 | CARD16 | TUBB | CCL4 | |
| TRABD2A | MAP3K8 | CD5 | ZBP1 | UQCRC1 | ABI3 | |
| TXK | IDI1 | ZNF683 | CD38 | ANXA5 | ARPC5L | |
| SOCS3 | CD74 | PRF1 | PLSCR1 | SLBP | APOBEC3G | |
| CD55 | JUN | EFHD2 | JAK2 | CDCA7 | LPCAT1 | |
| SATB1 | GABARAPL1 | ITGB1 | GBP1 | FABP5 | FCRL6 | |
| EEF1B2 | PMAIP1 | PLEK | OAS3 | DENND2D | DSTN | |
| PCED1B | TNFAIP3 | SPON2 | TYMP | PRDX3 | PTPN18 | |
| PIM2 | EML4 | FCGR3A | SAMD9 | SMC3 | CALR | |
| SESN3 | PPP2R5C | PRSS23 | SUB1 | DUT | FGR | |
| TPT1 | ISG20 | | LYST | NASP | CD3G | |
| MYC | SPOCK2 | | OAS2 | ANP32E | ADRB2 | |
| FHIT | ARL4C | | LDHA | CD81 | C1orf21 | |
| ABLIM1 | RSL24D1 | | SLFN5 | MKI67 | | |
| TNFSF8 | FYN | | PPM1K | MCM7 | | |
| SNX9 | MCL1 | | IFIT3 | TOX | | |
| SPINT2 | KLRB1 | | HSH2D | CXCR3 | | |
| EEF1A1 | COTL1 | | OASL | PCNA | | |
| PLP2 | | | GBP4 | MCM3 | | |
| NFKBIZ | | | | | | |
| INPP4B | | | | | | |
| EIF3E | | | | | | |
| NPM1 | | | | | | |
| SNHG8 | | | | | | |
| CD69 | | | | | | |
| VIM | | | | | | |
| EIF3L | | | | | | |

### S. Table 10

| Naïve-like | Tem | NKL/Temra | Exhaustion |
|---|---|---|---|
| CCR7 | CCL2 | GNLY | PDCD1 |
| LEF1 | CCL5 | ADGRG1 | LAG3 |
| IL7R | XCL2 | KLRD1 | HAVCR2 |
| LTB | XCL1 | KLRB1 | CTLA4 |
| SELL | CD69 | KIR2DL4 | HAVCR2 |
| TCF7 | GZMK | PRF1 | LAG3 |
| | IFNG | KLRC1 | CD160 |
| | | KLRG1 | BTLA |
| | | B3GAT1 | ENTPD1 |
| | | S1PR5 | |
| | | ZEB2 | |
| | | NKG7 | |
| | | FCRL6 | |
| | | KLRC2 | |
| | | NCAM1 | |
